# Prospective Registration of Trials: Where we are, why, and how we could get better

**DOI:** 10.1101/2024.07.24.24310935

**Authors:** Denis Mongin, Diana Buitrago-Garcia, Sami Capderou, Thomas Agoritsas, Cem Gabay, Delphine Sophie Courvoisier, Michele Iudici

**Author notes:** **Corresponding author:** Michele Iudici, Division of Rheumatology, Geneva University Hospitals and University of Geneva. Av. de Beau-Séjour 26, 1206 Genève.

## Abstract

**Objectives:** Transparent trial conduct requires prospective registration of a randomized controlled trial before the enrolment of the first participant. Registration aims to minimize potential biases through unjustified or hidden modification of trial design. We aimed to (1) estimate the proportion of randomized controlled trials that are prospectively registered and determine the time trends and the factors associated with prospective registration; (2) evaluate the reasons for non-adherence with prospective registration and explore potential mechanisms to enhance adherence with prospective registration. We studied trials published in rheumatology as a case study.

**Design and setting:** We searched for reports of trials in rheumatology published between January 2009 and December 2022 using MEDLINE-PubMed. We retrieved trial registration numbers using metadata and reviewed full texts. We conducted a multivariable logistic regression to identify factors associated with prospective trial registration. We sent an online survey to authors of trials that were not prospectively registered. We inquired about possible reasons for non-adherence with prospective registration and asked about potential solutions.

**Results:** We identified 1093 primary reports of randomized controlled trials; 453 (41.4%) were not prospectively registered. Of these, 130 (11.9%) were not registered, and 323 (29.5%) were retrospectively registered. Prospective registration increased over time at a rate of 3% per year (p<0.001), with only 13.3% (2/15) trials prospectively registered in 2009 to 73.2% (112/153) trials in 2022. Even among journals publicly supporting ICMJE recommendation, 16% of the trials published in 2022 were not prospectively registered. In the multivariable model, prospective registration was associated with a larger sample size, recruitment conducted across countries, and publication in a journal with a higher impact factor. Trial evaluating non pharmaceutical intervention, especially education, delivery of health care or wellness, had a lower rate of prospective registration. Investigators reported lack of knowledge, or organizational problems as the main reasons for retrospective registration. Authors also suggested linking ethical approval to trial registration as the best option to ensure prospective registration.

**Conclusions:** Despite significant improvement, adherence to prospective registration remains unsatisfactory in rheumatology. Different strategies targeting journal editors, healthcare professionals, and researchers may improve trial registration.

**Key findings:**

- Among 1093 randomized trials published in rheumatology between 2009 and 2022 30% were retrospectively registered and 12% not registered.
- Although the proportion of trials prospectively registered has increased over time, adherence remains suboptimal, even among trials published in journal endorsing ICMJE recommendations.
- Among the intervention tested by the trials, those concerned with education, delivery of health care or wellness had lower odds to be prospectively registered when compared to others.

What this adds to what is known related to methods research within the field of clinical epidemiology

- Although the amount of prospective registration has improved, our results raise the question about possible publication bias and other deleterious practices in research.

What is the implication, what should change now

- Solutions require attention at different levels, especially from researchers and journal editors. Prospective registration should be linked to the obtention of ethical approval, and reason for publishing trials not prospectively registered should be explicitly provided by the editors.

## INTRODUCTION

Randomized controlled trials (RCTs) are key drivers of clinical decision making and progress in medicine^1^. Ensuring transparent conduct and dissemination of RCTs findings provide reliable knowledge, maximize patient’s health and, avoid the publication of misleading results ^1,2^. One of the pillars of transparent research is the process by which the protocol of a RCT is made publicly available prospectively, before enrolling the first participant ^3^. Its aim includes the promotion of pre-planned analyses following a defined hypothesis, the minimization of unjustified and hidden modification of trial design during the study course ^4^, and the reduction of deleterious practices in science such as *p*-picking, HARKING (Hypothesizing After the Results are Known), switching outcomes or selective outcome reporting ^5,6^. Trial registration, particularly when informed by a systematic review also helps map research in a field, understand intervention trajectories, grasp research context, and prevent unnecessary duplication of research ^7^.

The Declaration of Helsinki ^8^, and the International Committee of Medical Journals Editors (ICMJE) support prospective registration ^3^. The World Health Organization (WHO) considers registering interventional studies a ‘*scientific, ethical, and moral responsibility*’ ^9^. To be published in journals that are members or support the ICMJE ^3^, RCTs that started recruiting patients after September 2005 are supposed to provide a prospective registration on a free web repository. National or international clinical trial registries have been increasingly established throughout the world, with WHO centralizing their information on a Clinical Trials Search Portal ^10^.

Despite these efforts, failure to adhere to prospective registration is still an issue in medicine ^11,12^. As an example, a study found that less than 15% of RCTs published in the five top psychiatry journals have been prospectively registered ^13^. Even in the top ranking journal of medicine, 28% of the published RCT have been found to be not prospectively registered ^11^. In rheumatology, a recent study ^16^ evidenced that only 66% of rheumatology journals required a registration to consider a RCT for publication, but the prevalence of adherence to prospective registration in the field is unknown.

Several studies have proposed various factors associated with lack of prospective registration ^14,15^, among which the journal impact factor, if the intervention is pharmaceutical or not, the funding scheme, or the number of country involved in the study. But to date, details on the time evolution of prospective registration, and on how this practice differs between types of interventions or on the reasons for the lack of adherence are still missing.

In this study, we propose to study Rheumatology as a case study to assess the trends of prospective registration, the factors associated with the nonadherence with prospective registration, and explored the reason for such non-adherence and possible solutions to overcome it.

## METHODS

### Eligibility criteria of included studies

We included primary reports of RCTs in rheumatology published between 2009 and 2022 that started enrolling participants after 2005. We defined a RCT as a clinical study randomly allocating participants to different interventions. We excluded secondary publications of RCTs (post-hoc analyses, long term extension, secondary or additional analysis of RCTs), non-randomized or quasi-randomized studies, editorials, letters, protocols, erratum, corrigendum, systematic reviews, and studies not conducted on humans.

### Search strategy of rheumatology RCT manuscripts

On February 1, 2023, using MEDLINE-PubMed and the strategy conducted by Al-Durra et al. ^4^, we searched all primary reports of RCTs published in the journals listed in the category “rheumatology” of the 2022 Journal Citation Reports of Clarivate ^17^, as well as all primary reports of rheumatology RCTs in the five top journals in general medicine (The Lancet, New England Journal of Medicine, JAMA, BMJ and Annals of Internal Medicine). Detailed search strategy and the list of included journals can be found in supplementary tables 1-2 and appendix1 - 2.

#### Selection of eligible manuscripts

Two authors reviewed titles and abstracts (SC and MI). If needed full texts were evaluated. Disagreements were resolved by a third reviewer (DM or DBG).

#### Data extraction

##### Identification of trial registration number

Presence of a Trial Registration Number (TRN) in each included study was determined automatically by first checking the PubMed-Medline metadata.^18^ After, the full text was reviewed using a list a regular expression patterns (Supplementary Appendix 3). If more than one TRN was found, one author (SC) checked each TRN against the registration data reported in the WHO-ICTRP database to identify the TRN corresponding to the publication. If no TRN was found in metadata or full text, the full text and supplementary material was reviewed. Also, an additional search on Google, Google Scholar and the Cochrane library was conducted. If no TRN was identified, the corresponding authors were contacted to inquiry about the TRN. If no TRN was found after these steps, the trial was considered *as not registered*.

##### Study outcome and definition of registration status

The primary outcome of the present study was the prospective registration of the RCT. A published RCT was considered prospectively registered if an associated TRN was found and had a registration date less than 30 days after the date of first recruitment.

#### Data Analysis

We analyzed the findings in two phases. First, we estimated the temporal trends of prospective registration of rheumatology trials and the factors associated with the failure of prospective RCT registration. Second, we investigated the reasons for non-prospective registration through an online survey sent to the contact authors of the trials that were not registered prospectively.

In the descriptive analyses, continuous variables were compared with t-test or Mann-Whitney test, and categorical variables with chi-square test or Fisher’s exact test, as appropriate. All statistical analyses were performed using R 4.2.0 and the xml2 library to handle xml files, the easyPubMed library to perform MEDLINE queries in PubMed and the rvest library to automatically recover data from websites.

##### Part 1: Prospective versus retrospective trial registration trends in rheumatology

Based on previous studies ^11,19,20^, we considered the following variables as potential factors related to registration practices: type of intervention, funding, sample size, countries of enrolment, year of the trial publication, journal impact factor, number of authors, and journal ICMJE status (for details on variables retrieval and operationalization, (Supplementary Appendix 4-5). The association between prospective trial registration and the chosen covariates was estimated using univariable and multivariable logistic regression. Linearity of the association between prospective trial registration and sample size and impact factor was assessed visually. We considered for both covariates a piecewise linear relation with prospective registration composed of two segments, and the knots were estimated using segmented modelling ^21^. Missing covariates were handled with multiple imputation with chained equation, using 20 samples and 5 iteration and using the outcome and the ensemble of covariates in the model. Estimates of regression were pooled according to Rubin’s law ^22.^.

##### Part 2: Reasons for noncompliance with prospective registration in rheumatology

An online survey was sent to the corresponding authors and/or to the designated trial contact persons of trials not registered prospectively. The questions used in our survey (Supplementary Appendix 6) were based on the survey published by Hunter et al. (17) and aimed at investigating the reasons for nonadherence with the prospective registration and explore the investigators’ opinion about possible measures that could have prevented this issue. The online survey was sent and stored using the electronic data capture system (REDCap, Vanderbilt University, Nashville, TN, USA) ^23^.

##### Ethics

The Geneva Research Ethics Committee Ethics committee exempted the present study from formal ethics review since we used publicly available data.

## RESULTS

### Identification of Trial Registration Number (TRN)

We screened 1,707 records and identified 1093 primary reports of RCTs, 41.4% (453/1023) of them were not registered prospectively. Among these 11.9% (130/1093), had been not registered and, 29.5% (323/1093) were retrospectively registered) (Supplementary Table 3).

#### Characteristics of the included trials

Most studies were conducted in a single country 79.7%, (872/1093) and were not industry-funded 57.8% (632/1093), The majority of included RCTs assessed pharmacological interventions 42.6% (466/1093 42.6) followed by interventions related to exercise therapy 13.9% (152/1093) and procedures 13.6% (149/1093). About half of the included studies were published in journals endorsing the ICMJE recommendations 51.4% (562/1093). From the included trials, 40.0% (421/1093), were funded by the industry and focused on pharmacological interventions 73.6% (310/466) (Table 1). Among RCTs published in journals endorsing ICMJE recommendations, 27.5% (155/562), were not prospectively registered, the number decreased in time to reach 16% (12/75) in 2022. In the overall sample, 21.7% (122/562) of the RCTs were retrospectively registered, this number also that decreased over time. Around5.9% (33/562) of them were not registered at all, a proportion that remained stable in time (Supplementary Table 4). Part 1: Prospective versus retrospective trial registration trends in rheumatology

**Table 1.**
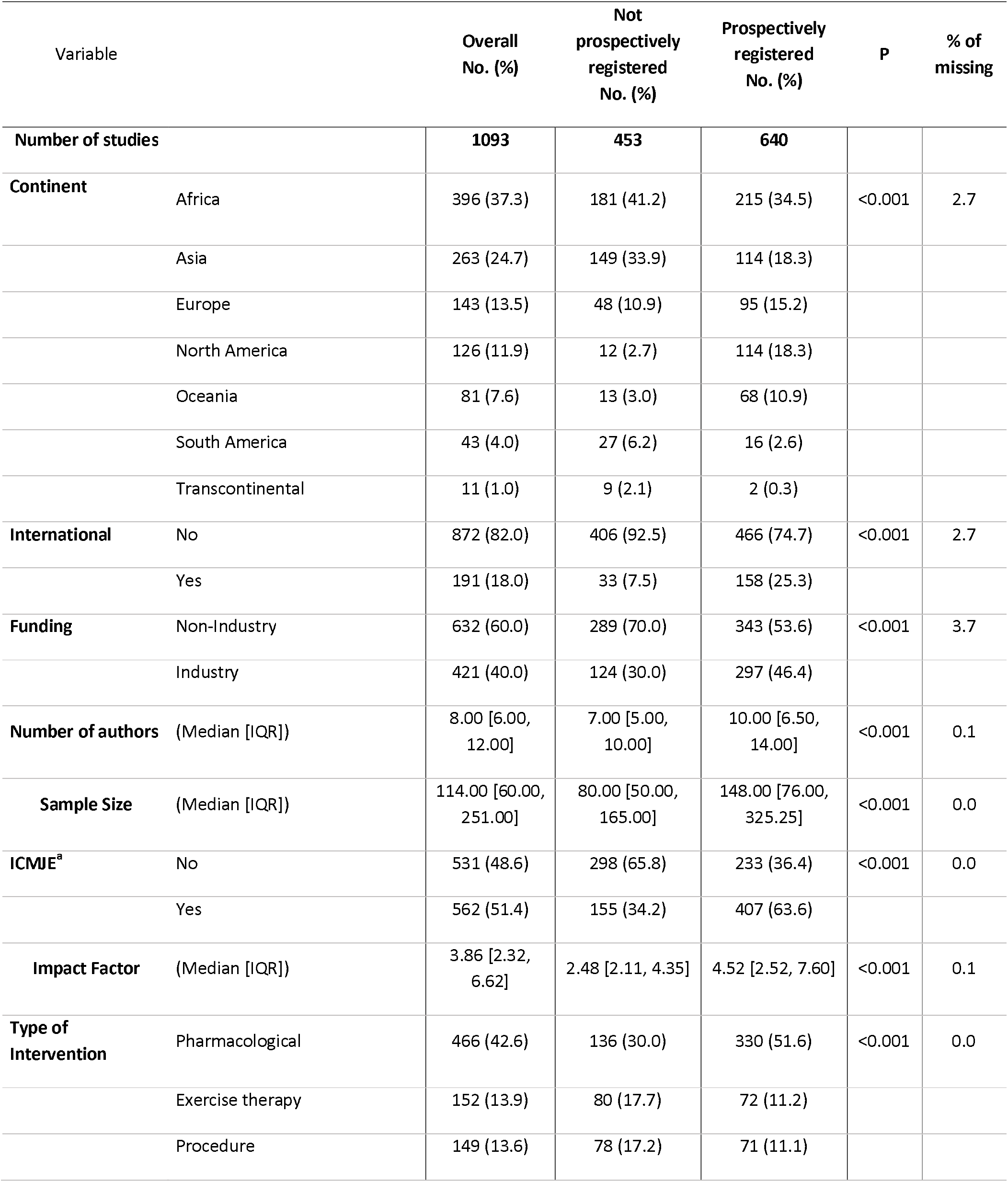
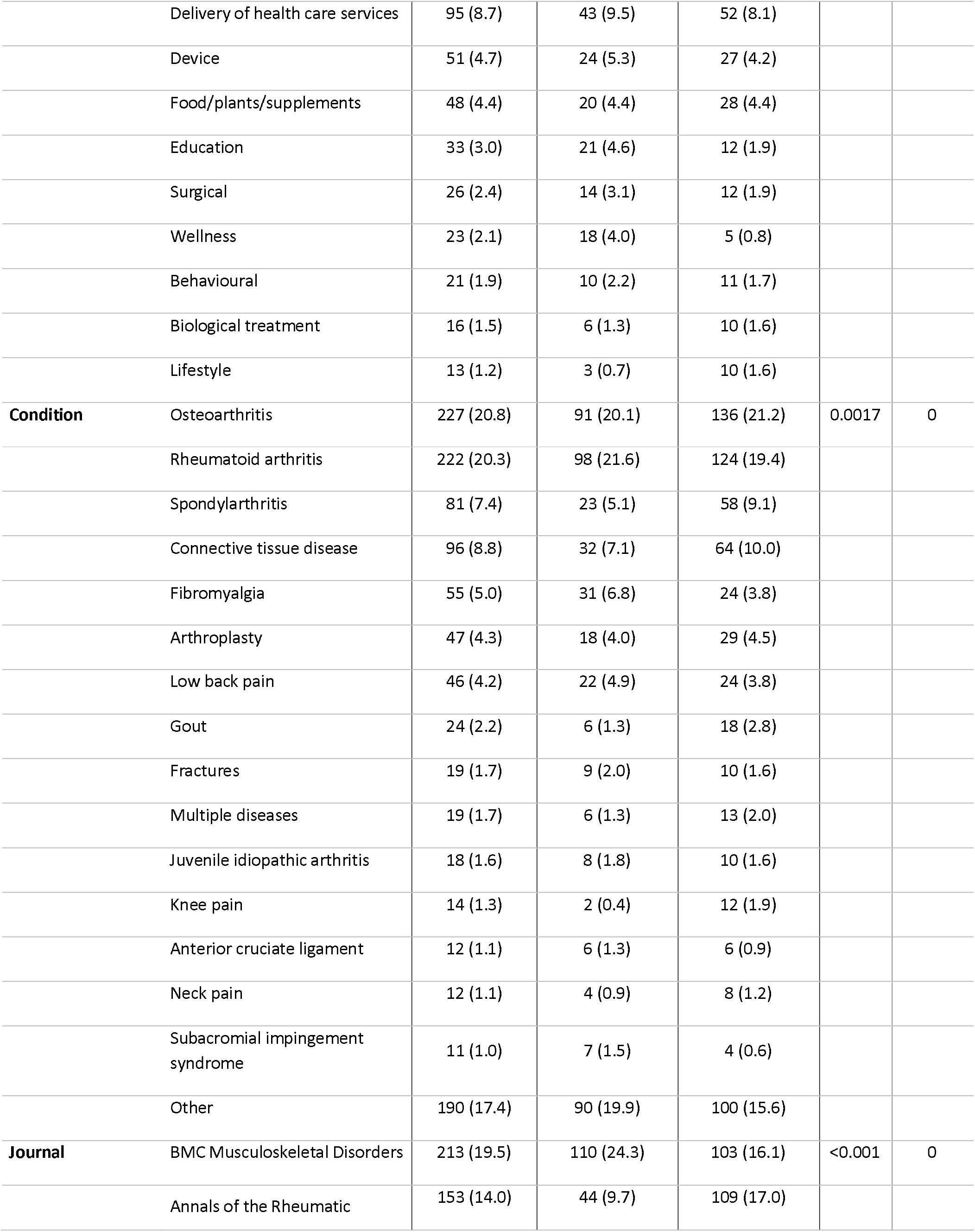
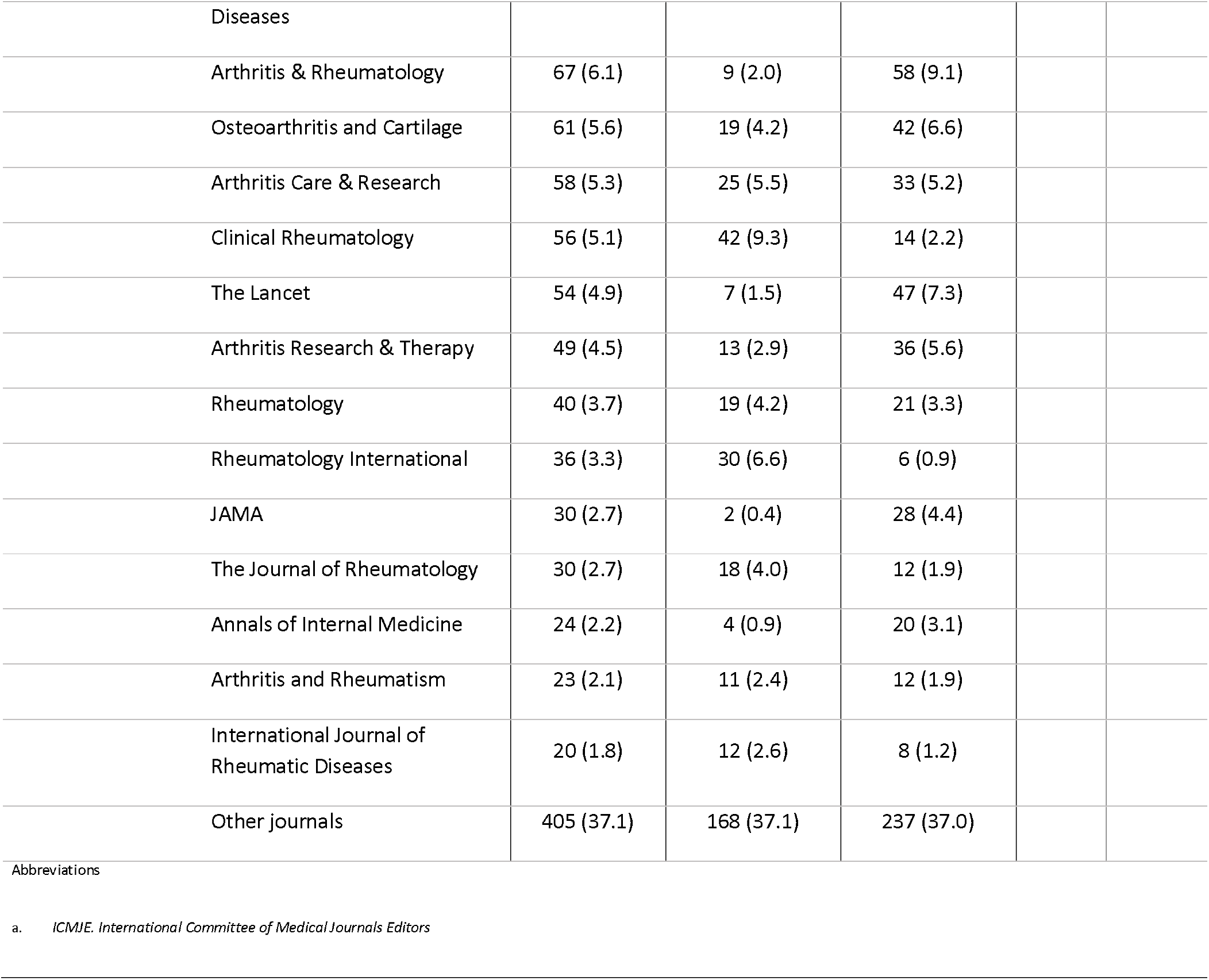
Characteristics of the included randomized controlled trials.

| Variable |  | Overall<br>No. (%) | Not<br>prospectively<br>registered<br>No. (%) | Prospectively<br>registered<br>No. (%) | P | % of<br>missing |
| --- | --- | --- | --- | --- | --- | --- |
| **Number of studies** |  | **1093** | **453** | **640** |  |  |
| **Continent** | Africa | 396 (37.3) | 181 (41.2) | 215 (34.5) | <0.001 | 2.7 |
|  | Asia | 263 (24.7) | 149 (33.9) | 114 (18.3) |  |  |
|  | Europe | 143 (13.5) | 48 (10.9) | 95 (15.2) |  |  |
|  | North America | 126 (11.9) | 12 (2.7) | 114 (18.3) |  |  |
|  | Oceania | 81 (7.6) | 13 (3.0) | 68 (10.9) |  |  |
|  | South America | 43 (4.0) | 27 (6.2) | 16 (2.6) |  |  |
|  | Transcontinental | 11 (1.0) | 9 (2.1) | 2 (0.3) |  |  |
| **International** | No | 872 (82.0) | 406 (92.5) | 466 (74.7) | <0.001 | 2.7 |
|  | Yes | 191 (18.0) | 33 (7.5) | 158 (25.3) |  |  |
| **Funding** | Non-Industry | 632 (60.0) | 289 (70.0) | 343 (53.6) | <0.001 | 3.7 |
|  | Industry | 421 (40.0) | 124 (30.0) | 297 (46.4) |  |  |
| **Number of authors** | (Median [IQR]) | 8.00 [6.00, 12.00] | 7.00 [5.00, 10.00] | 10.00 [6.50, 14.00] | <0.001 | 0.1 |
| **Sample Size** | (Median [IQR]) | 114.00 [60.00, 251.00] | 80.00 [50.00, 165.00] | 148.00 [76.00, 325.25] | <0.001 | 0.0 |
| **ICMJE<sup>a</sup>** | No | 531 (48.6) | 298 (65.8) | 233 (36.4) | <0.001 | 0.0 |
|  | Yes | 562 (51.4) | 155 (34.2) | 407 (63.6) |  |  |
| **Impact Factor** | (Median [IQR]) | 3.86 [2.32, 6.62] | 2.48 [2.11, 4.35] | 4.52 [2.52, 7.60] | <0.001 | 0.1 |
| **Type of Intervention** | Pharmacological | 466 (42.6) | 136 (30.0) | 330 (51.6) | <0.001 | 0.0 |
|  | Exercise therapy | 152 (13.9) | 80 (17.7) | 72 (11.2) |  |  |
|  | Procedure | 149 (13.6) | 78 (17.2) | 71 (11.1) |  |  |
|  | Delivery of health care services | 95 (8.7) | 43 (9.5) | 52 (8.1) |  |  |
|  | Device | 51 (4.7) | 24 (5.3) | 27 (4.2) |  |  |
|  | Food/plants/supplements | 48 (4.4) | 20 (4.4) | 28 (4.4) |  |  |
|  | Education | 33 (3.0) | 21 (4.6) | 12 (1.9) |  |  |
|  | Surgical | 26 (2.4) | 14 (3.1) | 12 (1.9) |  |  |
|  | Wellness | 23 (2.1) | 18 (4.0) | 5 (0.8) |  |  |
|  | Behavioural | 21 (1.9) | 10 (2.2) | 11 (1.7) |  |  |
|  | Biological treatment | 16 (1.5) | 6 (1.3) | 10 (1.6) |  |  |
|  | Lifestyle | 13 (1.2) | 3 (0.7) | 10 (1.6) |  |  |
| **Condition** | Osteoarthritis | 227 (20.8) | 91 (20.1) | 136 (21.2) | 0.0017 | 0 |
|  | Rheumatoid arthritis | 222 (20.3) | 98 (21.6) | 124 (19.4) |  |  |
|  | Spondylarthritis | 81 (7.4) | 23 (5.1) | 58 (9.1) |  |  |
|  | Connective tissue disease | 96 (8.8) | 32 (7.1) | 64 (10.0) |  |  |
|  | Fibromyalgia | 55 (5.0) | 31 (6.8) | 24 (3.8) |  |  |
|  | Arthroplasty | 47 (4.3) | 18 (4.0) | 29 (4.5) |  |  |
|  | Low back pain | 46 (4.2) | 22 (4.9) | 24 (3.8) |  |  |
|  | Gout | 24 (2.2) | 6 (1.3) | 18 (2.8) |  |  |
|  | Fractures | 19 (1.7) | 9 (2.0) | 10 (1.6) |  |  |
|  | Multiple diseases | 19 (1.7) | 6 (1.3) | 13 (2.0) |  |  |
|  | Juvenile idiopathic arthritis | 18 (1.6) | 8 (1.8) | 10 (1.6) |  |  |
|  | Knee pain | 14 (1.3) | 2 (0.4) | 12 (1.9) |  |  |
|  | Anterior cruciate ligament | 12 (1.1) | 6 (1.3) | 6 (0.9) |  |  |
|  | Neck pain | 12 (1.1) | 4 (0.9) | 8 (1.2) |  |  |
|  | Subacromial impingement syndrome | 11 (1.0) | 7 (1.5) | 4 (0.6) |  |  |
|  | Other | 190 (17.4) | 90 (19.9) | 100 (15.6) |  |  |
| **Journal** | BMC Musculoskeletal Disorders | 213 (19.5) | 110 (24.3) | 103 (16.1) | <0.001 | 0 |
|  | Annals of the Rheumatic | 153 (14.0) | 44 (9.7) | 109 (17.0) |  |  |

|  |  |  |  |
| --- | --- | --- | --- |
| Diseases |  |  |  |
| Arthritis & Rheumatology | 67 (6.1) | 9 (2.0) | 58 (9.1) |
| Osteoarthritis and Cartilage | 61 (5.6) | 19 (4.2) | 42 (6.6) |
| Arthritis Care & Research | 58 (5.3) | 25 (5.5) | 33 (5.2) |
| Clinical Rheumatology | 56 (5.1) | 42 (9.3) | 14 (2.2) |
| The Lancet | 54 (4.9) | 7 (1.5) | 47 (7.3) |
| Arthritis Research & Therapy | 49 (4.5) | 13 (2.9) | 36 (5.6) |
| Rheumatology | 40 (3.7) | 19 (4.2) | 21 (3.3) |
| Rheumatology International | 36 (3.3) | 30 (6.6) | 6 (0.9) |
| JAMA | 30 (2.7) | 2 (0.4) | 28 (4.4) |
| The Journal of Rheumatology | 30 (2.7) | 18 (4.0) | 12 (1.9) |
| Annals of Internal Medicine | 24 (2.2) | 4 (0.9) | 20 (3.1) |
| Arthritis and Rheumatism | 23 (2.1) | 11 (2.4) | 12 (1.9) |
| International Journal of Rheumatic Diseases | 20 (1.8) | 12 (2.6) | 8 (1.2) |
| Other journals | 405 (37.1) | 168 (37.1) | 237 (37.0) |

Abbreviations
a. ICMJE. *International Committee of Medical Journals Editors*

When compared to non-prospectively registered RCTs, prospectively registered RCTs were more often conducted in more than one country 92.5% (406/453) vs 74.7% (466/640), assessed more frequently pharmacological interventions 51.6% (330/640) vs 30.0% (136/453), among those registered prospectively 48.4% (310/640) were industry funded. Also, prospectively registered RCTs had a larger median sample size (148 vs 80 participants), had more authors (median 7 vs 10 authors), were more frequently published in ICMJE journals 63.6% (407/453) vs 34.2% (155/640) and, had a higher median impact factor (4.52 vs 2.48) (Table 1). The proportion of prospectively registered RCTs increased over time at a rate of three percentage points per year (p<0.001), with less than one-third of trials prospectively registered in 2009, against 73.2% in 2022 (Figure 2 and Supplementary Table 4).

**Figure 1.**
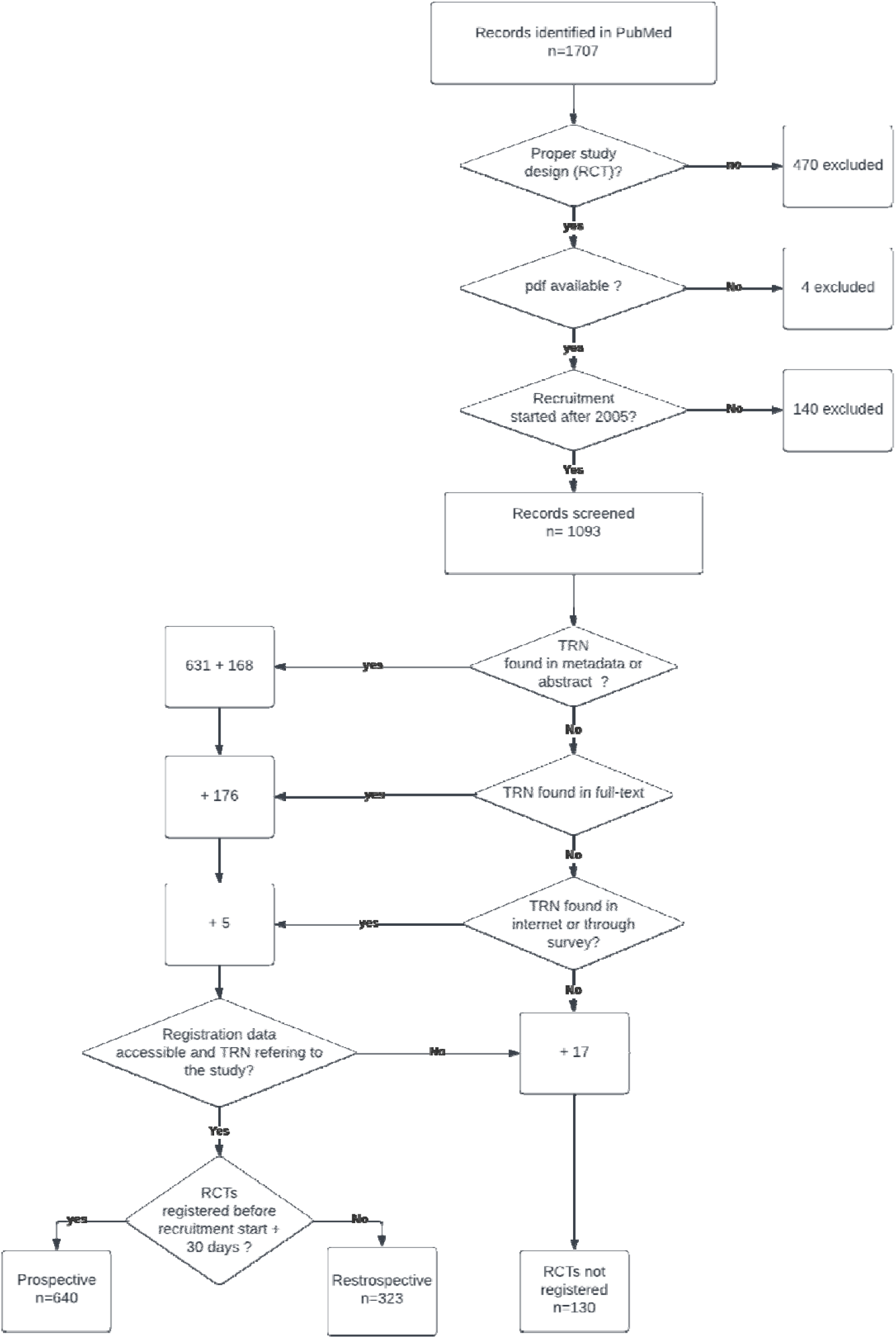
Flow diagram study selection. TRN search and retrieval of prospective/ retrospective status. Values correspond to the number of publications

**Figure 2.**
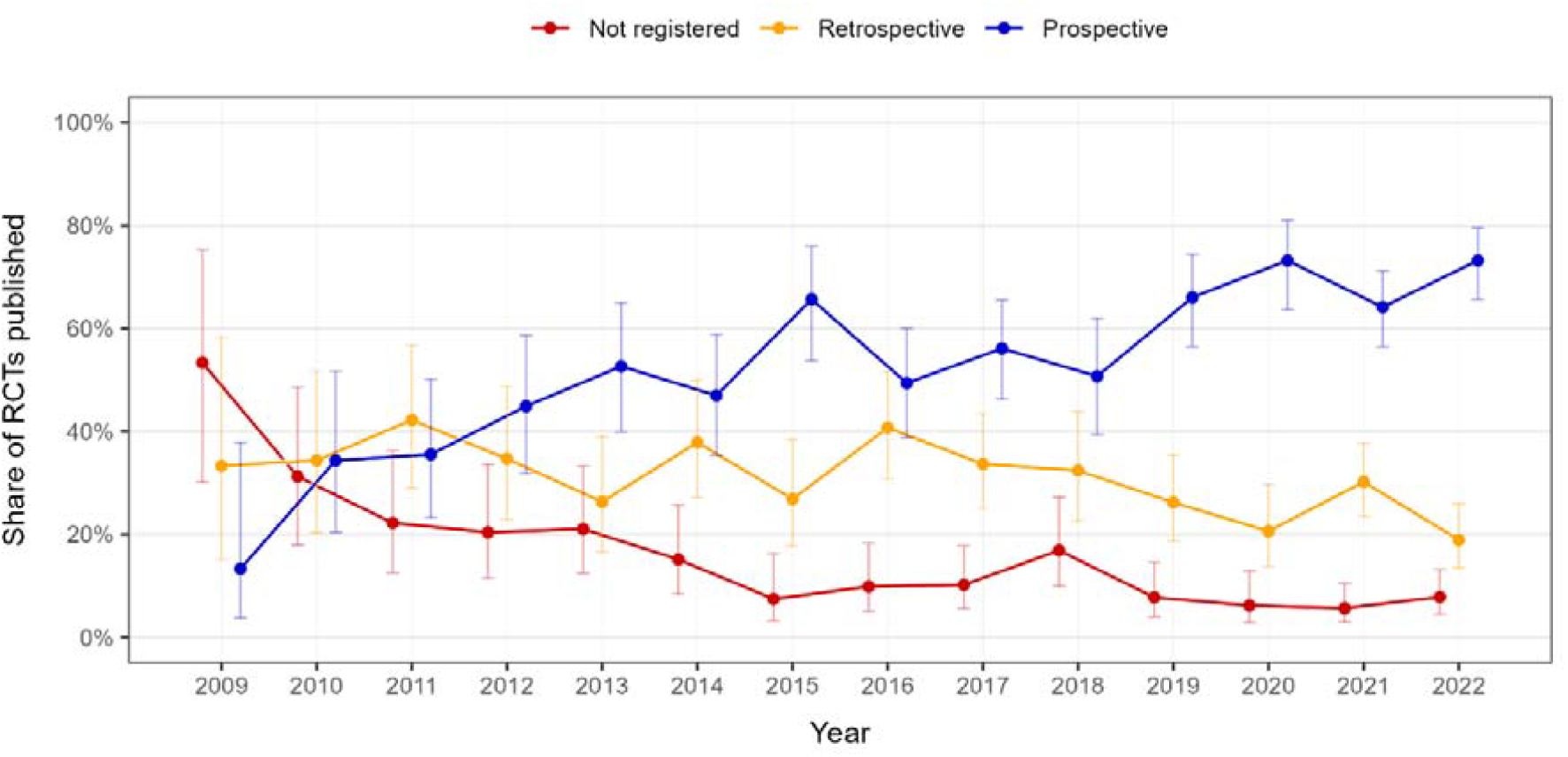
Time evolution proportion of prospectively registered, retrospectively registered and not registered published RCTs by year. Error bars are the confidence interval provided by the Wilson score interval

All these factors were significantly associated with prospective registration in the univariable analysis (Table 2). The association of prospective registration and either impact factor or sample size (Supplementary Figure1) showed a clear two pieces linear relationship, with a knot at 500 [95%CI: 108-892] for sample size and 5.5 [95%CI: 3.3-7.6] for impact factor. The sample size increased the odds of being prospectively registered when below 500 participants (OR 1.46 95%CI [1.32, 1.60] for every 100 participants) but had no effect on prospective registration when above 500 participants (OR 0.99 95%CI [0.98-1.02]). Similarly, the impact factor increased the odds of prospective registration when the impact factor was below 5.5, (OR 1.54 95%CI [1.39, 1.71]) but this association was weaker when the impact factor was above 5.5 (OR 1.10 95%CI [1.08, 1.12]).

**Table 2.** Factors associated with prospective trial registration in rheumatology trials.

| Characteristic | Univariable |  | Multivariable |  |
| --- | --- | --- | --- | --- |
|  | OR [95% CI] | P value | OR [95% CI] | P value |
| **International Trial** |  |  |  |  |
| No | Reference |  | Reference |  |
| Yes | 4.10 [2.76, 6.07] | <0.001 | 1.84 [1.12, 3.01] | 0.02 |
| **Funding** |  |  |  |  |
| Non-Industry | Reference |  | Reference |  |
| Industry | 2.02 [1.55, 2.63] | <0.001 | 1.11 [0.79, 1.56] | 0.53 |
| **Year** |  |  |  |  |
|  | 1.13 [1.09, 1.17] | <0.001 | 1.16 [1.11, 1.20] | <0.001 |
| **Number of authors** |  |  |  |  |
|  | 1.10 [1.07, 1.12] | <0.001 | 1.01 [0.98, 1.03] | 0.68 |
| **Journal endorsed ICMJE recommendations<sup>1</sup>** |  |  |  |  |
| No | Reference |  | Reference |  |
| Yes | 3.36 [2.61, 4.32] | <0.001 | 1.18 [0.78, 1.79] | 0.43 |
| **Intervention** |  |  |  |  |
| Non-pharmacological | Reference |  | Reference |  |
| Pharmacological | 2.50 [1.94, 3.22] | <0.001 | 1.40 [1.01, 1.95] | 0.047 |
| **Impact factor if < 5.5** |  |  |  |  |
|  | 1.54 [1.39, 1.71] | <0.001 | 1.38 [1.18, 1.61] | <0.001 |
| **Impact factor if > 5.5** |  |  |  |  |
|  | 1.10 [0.83, 1.45] |  | 1.03 [0.71, 1.49] |  |
| **Sample size if < 500, per 100 participants** |  |  |  |  |
|  | 1.46 [1.32, 1.60] | <0.001 | 1.21 [1.09, 1.35] | <0.001 |
| **Sample size if > 500, per 100 participants** |  |  |  |  |
|  | 0.99 [0.99, 1.01] | 0.02 | 1.00 [0.99, 1.00] | 0.03 |

In the adjusted model, the factors that remained independently and positively associated with prospective registration were recruitment was done across multiple countries(OR 1.84 95%CI [1.12, 3.01]), the impact factor when it was below 5.5 (OR 1.38 95%CI [1.18, 1.61]), the planned sample size when below 500 participants (OR 1.21 95%CI [1.09, 1.35]), and the year of publication, (OR of 1.16 95%CI [1.11, 1.20] per year).

Regarding interventions categories (eTable5), educational interventions (OR 0.64 95% CI [0.14, 0.79]), interventions associated to the delivery of health care (OR 0.57 95% CI [0.34, 0.97]), and wellness and spa interventions (OR 0.24 95% CI [0.08, 0.70]) significantly reduced the odds of prospective registration. Regarding health conditions, RCTs assessing interventions for knee pain, neck pain or low back pain (OR 14.14 [2.74, 73.06], 8.01 [2.06, 31.05], 3.56 [1.69, 7.50]), interventions for fractures (OR 3.51 [1.27, 9.70]), interventions related to arthroplasty (OR 3.11 [1.51, 6.39]) and to osteoarthritis (OR 1.99 [1.26, 3.15]) had a significantly higher probability to be prospectively registered when compared to RCTs in rheumatoid arthritis (Supplementary Table 6).

#### Part 2: Reasons for non-adherence with prospective registration in rheumatology

##### Retrospective registered trials

We sent 365 survey invitations (296 corresponding authors, 69 clinical trial contacts) and, 42 accepted to participate (response rate 11.5%). Compared to non-respondents, respondents were involved in studies more frequently published in journals endorsing ICMJE recommendations (47.6% vs. 32.9%), and with a higher impact factor (Supplementary Table 7). The main reasons for retrospective registration were a lack of knowledge (45.0%), either about the need for prospective registration in general or, less frequently, about the type of trial requiring prospective registration (Figure 3, left panel), and time constraints (17.0%). As far as potential factors that could prevent this issue, respondents suggested linking the obtention for Ethics approval to trial registration as the best option to ensure prospective registration (Figure 3, right panel).

**Figure 3.**
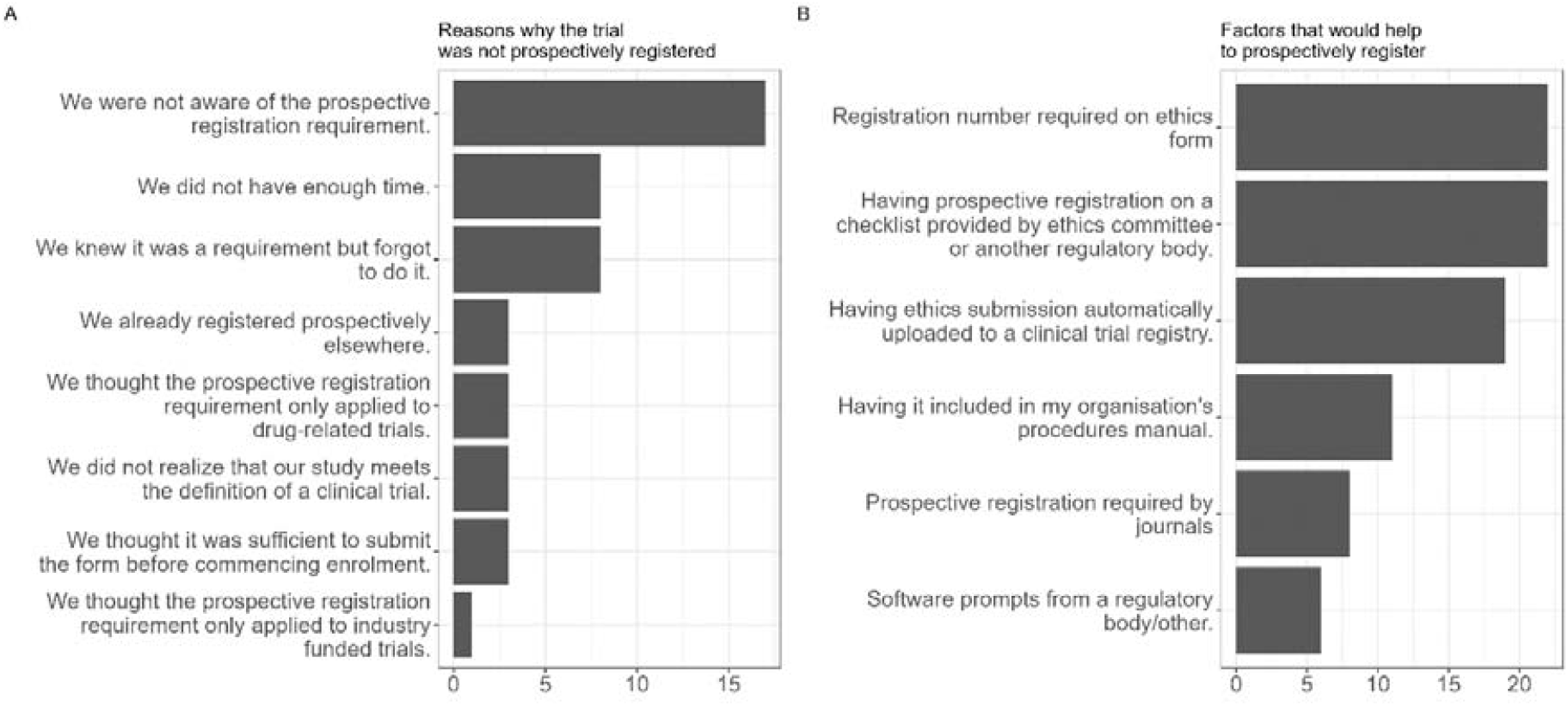
Survey results: Reasons for non-prospective trial registration and preventive actions. Reasons for retrospective registration (left) and factors that could prevent it (right). Answers were chosen from a list of options. Free-text answers were recategorized among existing answers

##### Unregistered trials

We sent 126 survey invitations to contact authors and only 14 authors accepted to participate (response rate 11.1%). Although most of the characteristics of the trials were similar among respondents and non-respondents, a higher proportion of non-respondents published their RCT results in journals endorsing ICMJE recommendations (75.9% vs. 50%) (eTable8). The main reason for non-registration was lack of awareness (6 out of 7 responses). Similarly, respondents suggested having the registration as part of the requirements for Ethics approval or a mandatory requirement from scientific journals to ensure prospective trial registration.

## DISCUSSION

Our study included over 1,000 RCTs published in rheumatology journals from 2009 to 2022. Of these 41.4% were not registered prospectively, with 29.5% retrospectively registered, and 11.9 % not registered at all. Over time, registration practices gradually improved. The factors associated with better trial registration were recruitment conducted across countries, a higher sample size among small trials (below 500 participants), publication in a journal with a higher impact factor and trials investigating pharmacological interventions. The lower odds of prospective registration among non-pharmacological interventions were mainly caused by intervention concerned with education, delivery of health care and wellness.

Despite efforts supporting trial registration, and although registration of trials is mandatory since 2005 in the United States and Europe, adherence to prospective trial registration remains inadequate in rheumatology. The gradual improvement in registration is slow, making adherence an unsolved issue in medicine. Adherence to prospective trial registration in the Australian New Zealand Clinical Trials Registry (ANZCTR) was 48% in 2006 and 64% in 2015^19^. Lower rates of prospective trial registration have been reported in a study analyzing a large sample of clinical trials included in 100 Cochrane reviews (only 31% were registered, of which just one-third prospectively) ^15^ in surgical trials 50% ^24^, and in trials investigating drugs for the prevention of postoperative nausea and vomiting 36%^25^. Also, a large study analyzing the registration status of more than 10’000 trials found adherence with prospective registration in 42% of cases ^4^. Comparing our findings with those from other medical specialties is challenging due to variations in study eligibility criteria, investigation periods, and the definition of prospective registration ^26^. We considered a trial as prospectively registered if an associated TRN was found with a registration date less than 30 days after the date of first recruitment, to incorporate also the FDA regulation that requires all submitted trials to be registered no later than 21 days after enrolment ^27^, and to take into account those trials whose registration/start date were provided at month level. However, when other studies applied a more conservative definition of prospective registration (as we did), the observed adherence to registration requirements were comparable to our results ^28^. Altogether, these findings underline that a high proportion of trials, whose results drive daily practice, may suffer from deleterious and difficult to identify and amend issues such as publication bias, selective outcome reporting, or data-driven analysis. These practices are associated with the risk of yielding false-positive results, potentially leading to the implementation of ineffective or harmful measures^29^ and, finding effective solutions to this issue is a paramount concern for public health.

Though adherence to prospective registration among trials published in journals endorsing ICMJE guidelines was considerably higher, 16% of RCTs were still accepted without prospective registration, highlighting that some journals do not consistently adhere to their registration policies. The reasons for accepting and publishing unregistered or retrospectively registered trials might be related to hesitance to penalize valid research or studies from developing countries, concerns about losing submissions to competing journals, or misunderstandings about the relevance of registration policies ^30^. Although this is rarely disclosed, it would be relevant that journals accepting trials that either did not register or did it retrospectively state their reason for accepting them ^25,31^.

The reasons for non-prospective registration provided by the authors who answered our survey were consistent with other studies. Reveiz et al. ^32^, two years after the publication of ICMJE recommendations on trial registration, found that one-third of the researchers identified the lack of knowledge as the predominant factor for not registering trials prospectively. More recently, a survey conducted by Ranawaka et al. ^33^ among participants in a medical congress, including active researchers, further highlighted suboptimal awareness regarding the necessity and perceived importance of prospective trial registration. Developing a procedure enabling researchers to register RCTs prospectively as part of the submission to the Ethics Committee appears as a potential solution, already implemented in some countries ^19^. Besides the lack of awareness about registration, other factors such as the time needed to register, cost and complexity of registration, and generic ‘organizational’ problems have also been identified as significant barriers to registration ^19,34^. Strategy prioritizing user-friendliness, speed, and affordability of the registration process may help increase the rate of prospective registration. Further measures may include legal frameworks and active enforcement of a prospective registration by funding bodies, ethical committees, and institutions, and of course the implementation of teaching modules emphasizing the relevance of study registration during medical and healthcare curricula.

### Limitations and strengths of this study

Some limitations should be acknowledged. First, we did not review the full text of RCTs to identify the reasons for retrospective or non-registration. However, we consider this a minor concern given that a recent study showed that only 3.5% of RCTs report the reasons for retrospective registration in the published manuscript ^35^. Second, there is a risk of selection bias due to the low proportion of participants in our survey that intended to determine the reasons for non-prospective registration, as respondents and non-respondents might have different profiles.

The strengths of our study include the broad time interval considered together with the amount of information used in the analysis, which makes it one of the most complete studies on the subject and the first one to address this relevant issue in Rheumatology. Also, the direct feedback from investigators about reasons for lack of registration or retrospective registration and their opinions on potential solutions provides better qualitative insights on adherence to registration.

## Conclusion

Despite the observed improvement over time, adherence with legal and ethical obligations of prospectively registering trials is still suboptimal in rheumatology. Even journals endorsing the ICMJE recommendations on registration practices still publish unregistered or retrospectively registered studies. As a result, a high proportion of trials whose results drive daily practice are likely to suffer from deleterious and difficult to identify issues such as publication bias, selective outcome reporting, or data-driven analysis. All these practices may contribute to spurious inferences about current best care, potentially leading to the implementation of ineffective or harmful measures (24). Our results strongly support the need to for implement educational and regulatory strategies to prevent these issues.

## Data Availability

Code and data are available at the following Gitlab repository:

https://gitlab.unige.ch/trial_integrity/prospective_registration_public

## Author Contributions

**Denis MONGIN:** Conceptualization, Methodology, Software, Formal analysis, Investigation, Data Curation, Funding acquisition, Writing - Original Draft, Visualization, Supervision

**Diana BUITRAGO-GARCIA:** Validation, Investigation, Data Curation, Writing - Review & Editing

**Sami CAPDEROU**: Formal analysis, Investigation, Data Curation, Writing - Original Draft, Software

**Thomas AGORITSAS**: Writing - Review & Editing, Conceptualization.

**Cem GABAY:** Writing - Review & Editing, Conceptualization.

**Delphine Sophie COURVOISIER:** Conceptualization, Methodology, Writing - Review & Editing

**Michele IUDICI:** Conceptualization, Methodology, Data Curation, Writing - Review & Editing, Supervision, Project administration, Funding acquisition

## Data sharing statement

Code and data are available at the following Gitlab repository: https://gitlab.unige.ch/trial_integrity/prospective_registration_public

## References

1. Moher D, Hopewell S, Schulz KF, et al. CONSORT 2010 explanation and elaboration: updated guidelines for reporting parallel group randomised trials. BMJ 2010;340:c869. 10.1136/bmj.c869

2. Ioannidis JPA. Hundreds of thousands of zombie randomised trials circulate among us. Anaesthesia 2021;76(4):444–7. 10.1111/anae.15297

3. Angelis CD, Drazen JM, Frizelle FA, et al. Clinical trial registration: a statement from the International Committee of Medical Journal Editors. The Lancet 2004;364(9438):911–2. 10.1016/S0140-6736(04)17034-7

4. Al-Durra M, Nolan RP, Seto E, Cafazzo JA. Prospective registration and reporting of trial number in randomised clinical trials: global cross sectional study of the adoption of ICMJE and Declaration of Helsinki recommendations. BMJ 2020;369.

5. Erasmus A, Holman B, Ioannidis JP. Data-dredging bias. BMJ Evid-Based Med 2022;27(4):209–11.

6. Chan A-W, Hróbjartsson A, Haahr MT, Gøtzsche PC, Altman DG. Empirical Evidence for Selective Reporting of Outcomes in Randomized Trials Comparison of Protocols to Published Articles. JAMA 2004;291(20):2457–65. 10.1001/jama.291.20.2457. 10.1001/jama.291.20.2457

7. Viergever RF, Li K. Trends in global clinical trial registration: an analysis of numbers of registered clinical trials in different parts of the world from 2004 to 2013. BMJ Open 2015;5(9):e008932.

8. WMA - The World Medical Association-WMA Declaration of Helsinki – Ethical Principles for Medical Research Involving Human Subjects. https://www.wma.net/policies-post/wma-declaration-of-helsinki-ethical-principles-for-medical-research-involving-human-subjects/.

9. World Health Organization-WHO. Clinical Trials, Questions and Answers. https://www.who.int/news-room/questions-and-answers/item/clinical-trials.

10. World Health Organization-WHO. International Clinical Trials Registry Platform (ICTRP). Accessed Mar 6, 2024. https://www.who.int/clinical-trials-registry-platform.

11. Dal-Ré R, Ross JS, Marušić A. Compliance with prospective trial registration guidance remained low in high-impact journals and has implications for primary end point reporting. J Clin Epidemiol 2016;75:100–7.

12. Tan AC, Jiang I, Askie L, Hunter K, Simes RJ, Seidler AL. Prevalence of trial registration varies by study characteristics and risk of bias. J Clin Epidemiol 2019;113:64–74. 10.1016/j.jclinepi.2019.05.009

13. Scott A, Rucklidge JJ, Mulder RT. Is mandatory prospective trial registration working to prevent publication of unregistered trials and selective outcome reporting? An observational study of five psychiatry journals that mandate prospective clinical trial registration. PloS One 2015;10(8):e0133718.

14. Al-Durra M, Nolan RP, Seto E, Cafazzo JA. Prospective registration and reporting of trial number in randomised clinical trials: global cross sectional study of the adoption of ICMJE and Declaration of Helsinki recommendations. bmj 2020;369.

15. Lindsley K, Fusco N, Li T, Scholten R, Hooft L. Clinical trial registration was associated with lower risk of bias compared with non-registered trials among trials included in systematic reviews. J Clin Epidemiol 2022;145:164–73. 10.1016/j.jclinepi.2022.01.012

16. Jelinek T, Shumard A, Modi J, et al. Endorsement of reporting guidelines and clinical trial registration across Scopus-indexed rheumatology journals: a cross-sectional analysis. Rheumatol Int 2023; 10.1007/s00296-023-05474-4

17. Journal Citation Reports - Home. Accessed Mar 11, 2024. https://jcr.clarivate.com/jcr/home.

18. MEDLINE®PubMed® XML Element Descriptions and their Attributes. Accessed Mar 13, 2024. https://www.nlm.nih.gov/bsd/licensee/elements_descriptions.html.

19. Hunter KE, Seidler AL, Askie LM. Prospective registration trends, reasons for retrospective registration and mechanisms to increase prospective registration compliance: descriptive analysis and survey. BMJ Open 2018;8(3):e019983.

20. Gopal AD, Wallach JD, Aminawung JA, et al. Adherence to the International Committee of Medical Journal Editors’(ICMJE) prospective registration policy and implications for outcome integrity: a cross-sectional analysis of trials published in high-impact specialty society journals. Trials 2018;19:1–13.

21. Muggeo VMR. Estimating regression models with unknown break-points. Stat Med 2003;22(19):3055–71. https://onlinelibrary.wiley.com/doi/abs/10.1002/sim.1545. 10.1002/sim.1545

22. Schafer JL, Olsen MK. Multiple Imputation for Multivariate Missing-Data Problems: A Data Analyst’s Perspective. Multivar Behav Res 1998;33(4):545–71. 10.1207/s15327906mbr3304_5. 10.1207/s15327906mbr3304_5

23. Harris PA, Taylor R, Thielke R, Payne J, Gonzalez N, Conde JG. Research electronic data capture (REDCap)—A metadata-driven methodology and workflow process for providing translational research informatics support. J Biomed Inform 2009;42(2):377–81. https://www.sciencedirect.com/science/article/pii/S1532046408001226.10.1016/j.jbi.2008.08.010

24. Hardt JL, Metzendorf M-I, Meerpohl JJ. Surgical trials and trial registers: a cross-sectional study of randomized controlled trials published in journals requiring trial registration in the author instructions. Trials 2013;14:1–9.

25. Riemer M, Kranke P, Helf A, et al. Trial registration and selective outcome reporting in 585 clinical trials investigating drugs for prevention of postoperative nausea and vomiting. BMC Anesthesiol 2021;21(1):1–10.

26. Trinquart L, Dunn AG, Bourgeois FT. Registration of published randomized trials: a systematic review and meta-analysis. BMC Med 2018;16:1–13.

27. FDAAA 801 and the Final Rule | ClinicalTrials.gov. Accessed Apr 24, 2024. https://clinicaltrials.gov/policy/fdaaa-801-final-rule.

28. Zarin DA, Tse T, Williams RJ, Rajakannan T. Update on trial registration 11 years after the ICMJE policy was established. N Engl J Med 2017;376(4):383–91.

29. Joober R, Schmitz N, Annable L, Boksa P. Publication bias: what are the challenges and can they be overcome? J Psychiatry Neurosci JPN 2012;37(3):149–52. 10.1503/jpn.120065

30. Wager E, Williams P. “Hardly worth the effort”? Medical journals’ policies and their editors’ and publishers’ views on trial registration and publication bias: quantitative and qualitative study. BMJ 2013;347:f5248. http://www.bmj.com/content/347/bmj.f5248.abstract.10.1136/bmj.f5248

31. ICMJE | Home. Accessed Mar 13, 2024. https://www.icmje.org/.

32. Reveiz L, Krleža-Jerić K, Chan A-W, De Aguiar S. Do trialists endorse clinical trial registration? Survey of a Pubmed sample. Trials 2007;8(1):30. 10.1186/1745-6215-8-30. 10.1186/1745-6215-8-30

33. Ranawaka UK, de Abrew A, Wimalachandra M, Wanigatunge CA, Rajapakse LC, Goonaratna C. Awareness of clinical trial registration among healthcare professionals: An observational study. J Evid-Based Med 2018;11(4):227–32. 10.1111/jebm.12327

34. Agha RA, Jafree DJ, Vella-Baldacchino M, et al. Surveying opinions of 149 registrants to the Research Registry: Awareness of and attitudes towards research registration. Int J Surg Lond Engl 2017;39:182–7. 10.1016/j.ijsu.2016.12.040

35. Haslberger M, Gestrich S, Strech D. Reporting of retrospective registration in clinical trial publications: a cross-sectional study of German trials. BMJ Open 2023;13(4):e069553. http://bmjopen.bmj.com/content/13/4/e069553.abstract.10.1136/bmjopen-2022-069553

