## Supplementary material for "Prospective Registration of Trials: Where we are, why, and how we could get better": supp_files

**Supplementary Table 1.List of rheumatology journals Included in the study**

| |
|---|
| Acta Reumatológica Portuguesa |
| Advances In Rheumatology |
| Aktuelle Rheumatologie |
| Annals of the Rheumatic Diseases |
| Archives Of Rheumatology |
| BMC Musculoskeletal Disorders |
| Clinical And Experimental Rheumatology |
| Clinical Rheumatology |
| Current Opinion in Rheumatology |
| Current Rheumatology Reports |
| Current Rheumatology Reviews |
| Egyptian Rheumatologist |
| European Journal of Rheumatology |
| Indian Journal of Rheumatology |
| International Journal of Rheumatic Diseases |
| International Journal of Rheumatology |
| Journal Of Clinical Rheumatology |
| Joint Bone Spine |
| Journal Of Rheumatic Diseases |
| Journal Of Rheumatology |
| Journal Of Scleroderma and Related Disorders |
| Lancet Rheumatology |
| Lupus |
| Lupus Science & Medicine |
| Modern Rheumatology |
| Musculoskeletal Care |
| Nature Reviews Rheumatology |
| Open Access Rheumatology-Research and Reviews |
| Osteoarthritis And Cartilage |
| Pediatric Rheumatology |
| Arthritis Care & Research |
| Best Practice and Research In Clinical Rheumatology |
| Reumatismo |
| Reumatologia Clinica |
| Revista Cubana De Reumatología |
| Rheumatic Disease Clinics of North America |
| Arthritis And Rheumatology |
| Rheumatology |
| Rheumatology Advances in Practice |
| Rheumatology And Therapy |
| Rheumatology International |
| RMD Open |

| |
|---|
| Scandinavian Journal of Rheumatology |
| Seminars In Arthritis and Rheumatism |
| Therapeutic Advances in Musculoskeletal Disease |
| Arthritis Research and Therapy |
| Turk Osteoporoz Dergisi |
| Zeitschrift Fur Rheumatologie |
| Arthritis & Rheumatism |

**Supplementary Table 2. List of included journals in general medicine**

| |
|---|
| The Lancet |
| New England Journal of Medicine |
| JAMA |
| BMJ |
| Annals of Internal Medicine |

**Supplementary Appendix 1. Search strategy for rheumatology journals**

"(0303-464X[IS] OR 2523-3106[IS] OR 0341-051X[IS] OR 0003-4967[IS] OR 2148-5046[IS] OR 1471-2474[IS] OR 0392-856X[IS] OR 0770-3198[IS] OR 1040-8711[IS] OR 1523-3774[IS] OR 1573-3971[IS] OR 1110-1164[IS] OR 2147-9720[IS] OR 0973-3698[IS] OR 1756-1841[IS] OR 1687-9260[IS] OR 1076-1608[IS] OR 1297-319X[IS] OR 2093-940X[IS] OR 0315-162X[IS] OR 2397-1983[IS] OR 2665-9913[IS] OR 0961-2033[IS] OR 2053-8790[IS] OR 1439-7595[IS] OR 1478-2189[IS] OR 1759-4790[IS] OR 1179-156X[IS] OR 1063-4584[IS] OR 1546-0096[IS] OR 2151-464X[IS] OR 1532-1770[IS] OR 0048-7449[IS] OR 1699-258X[IS] OR 1606-5581[IS] OR 0889-857X[IS] OR 2326-5191[IS] OR 1462-0324[IS] OR 2514-1775[IS] OR 2198-6576[IS] OR 0172-8172[IS] OR 2056-5933[IS] OR 0300-9742[IS] OR 0049-0172[IS] OR 1759-720X[IS] OR 1478-6354[IS] OR 2147-2653[IS] OR 0340-1855[IS] OR 0004-3591[IS]) AND 2009:2022 [DP] AND (trial[TI] OR rct[TI]) AND (randomized[TI] OR randomised[TI]) NOT (non-randomized[TI] OR non-randomised[TI] OR nonrandomized[TI] OR nonrandomised[TI]) NOT protocol[TI] NOT secondary analysis[TI] NOT (systematic-review[TI] OR meta-analysis[TI]) NOT (design[TI] AND methodology[TI])"

**Supplementary Appendix 2. Search strategy for general medicine journals**

"(1474-547X[IS] OR 1533-4406[IS] OR 1538-3598[IS] OR 1756-1833[IS] OR 1539-3704[IS]) AND 2009:2022 [DP] AND (trial[TI] OR rct[TI]) AND (randomized[TI] OR randomised[TI]) NOT (non-randomized[TI] OR non-randomised[TI] OR nonrandomized[TI] OR nonrandomised[TI]) NOT protocol[TI] NOT secondary analysis[TI] NOT (systematic-review[TI] OR meta-analysis[TI]) NOT (design[TI] AND methodology[TI]) AND (Vasculitis[Mesh] OR Polychondritis, Relapsing[Mesh] OR Dermatomyositis[Mesh] OR Lupus Erythematosus, Systemic[Mesh] OR Mixed Connective Tissue Disease[Mesh] OR Rheumatic Diseases[Mesh] OR Polymyositis[Mesh] OR Arthritis, Rheumatoid[Mesh] OR Spondylarthritis[Mesh] OR Osteoarthritis[Mesh] OR Crystal Arthropathies[Mesh] OR gout[tiab] OR chondrocalc*[tiab] OR Wegener[tiab] OR churg-strauss[tiab] OR granulomatosis[tiab] OR polyangiitis[tiab] OR *angiitis[tiab] OR polyarteritis[tiab] OR Takayasu[tiab] OR Horton[tiab] OR aortitis[tiab] OR giant cell[tiab] OR polymyalgia[tiab] OR scleroderma[tiab] OR systemic sclerosis[tiab] OR lupus[tiab] OR sjogren[tiab] OR dermatomyositis[tiab] OR polymyositis[tiab] OR undifferentiated connective tissue[tiab] OR UCTD[tiab] OR mixed connective tissue[tiab] OR MCTD[tiab] OR behcet[tiab] OR cogan[tiab] OR Schoenlein-Henoch[tiab] OR rheumatoid[tiab] OR Still disease[tiab] OR osteoarthr*[tiab] OR spondyl*[tiab] OR reactive arthritis[tiab] OR psoriatic arthritis[tiab] OR arthrose[tiab] OR arthritis[tiab])"

**Supplementary Appendix 3. Regex codes to retrieve TRN during the data extraction phase**

The list of regex codes (Full regex code in the table below) was build using an extensive list of TRNs retrieved from the metadata of the complete PubMed citation record of 2022. Shorter regex codes were also created to allow identifying ambiguous text patterns (e.g. misspelled TRN). If a potential ambiguous TRN was detected and not complete TRN was identified, the result was manually checked by two authors (SC, DM) for validation.

The TRN regex detection algorithm was tested against a list of 24,380 known pattern extracts from the metadata of the complete PubMed citation record of 2022 and yielded a sensitivity of 99,1%. The TRN not detected during this preliminary test had all syntax errors and were identified by the short regex code.

<table>
<tr><th>Registry Name</th><th>Databank</th><th>Full regex code</th><th>Short regex code</th></tr>
<tr><td>Australian New Zealand Clinical Trials Registry</td><td>ANZCTR</td><td>\bA[NZ]{0,}CTRN{0,}[,-:]{0,}\s{0,}[0-9]{12,}</td><td>ACTR</td></tr>
<tr><td>Brazilian Clinical Trials Registry</td><td>ReBec</td><td>RBR.{0,1}[[A-z][0-9]]{5,6}</td><td>RBR</td></tr>
<tr><td>Clinical Research Information Service - South Korea</td><td>CRiS</td><td>KCT\-{0,}[0-9]{7}</td><td>KCT</td></tr>
<tr><td>Clinicaltrials.gov</td><td>ClinicalTrials.gov</td><td>NCT[0-9]{8}</td><td>(?<![A-z])NCT</td></tr>
<tr><td>Clinical Trials Registry - India</td><td>CTRI</td><td>CTRI[/[0-9]]{13,}</td><td>IRCT</td></tr>
<tr><td>Clinical Trials Registry - India</td><td>CTRI</td><td>[0-9]{4}/[0-9]{2}/[0-9]{6}</td><td>IRCT</td></tr>
<tr><td>Cuban Public Registry of Clinical Trials</td><td>RPCEC</td><td>RPCEC[0-9]{8}</td><td>RPCE</td></tr>
<tr><td>EU Clinical Trials Register</td><td>EudraCT</td><td>EudraCT(-|[0-9]){3,}</td><td>Eudra</td></tr>
<tr><td>EU Clinical Trials Register</td><td>EudraCT</td><td>EUDRACT(-|[0-9]){3,}</td><td>EUDRA</td></tr>
<tr><td>EU Clinical Trials Register</td><td>EudraCT</td><td>EUCTR(-|[0-9]){3,}</td><td>EUCT</td></tr>
<tr><td>EU Clinical Trials Register</td><td>EudraCT</td><td>(?<![-])\b[0-9]{4}-A{0,1}[0-9]{5,6}.{0,1}[0-9]{2}</td><td>EudraCT</td></tr>
<tr><td>German Clinical Trials Register</td><td>DRKS</td><td>DRKS[0-9]{8}</td><td>DRKS</td></tr>
<tr><td>Iranian Registry of Clinical Trials</td><td>IRCT</td><td>IRCT[[A-z]|[0-9]]{12,15}</td><td>IRCT</td></tr>
<tr><td>International Standard Randomised Controlled Trial Number</td><td>ISRCTN</td><td>ISRCTN\s{0,1}:{0,1}[0-9]{7,8}</td><td>ISRC</td></tr>
<tr><td>Japan Primary Registries Network</td><td>JPRN</td><td>JPRN(-|[0-9]|[A-z]){8,}</td><td>JPRN</td></tr>
<tr><td>Lebanese Clinical Trials Registry</td><td>LBCTR</td><td>LBCTR</td><td>LBCT</td></tr>
<tr><td>Thai Clinical Trials Registry</td><td>TCTR</td><td>TCTR\s{0,1}[0-9]{11}</td><td>TCTR</td></tr>
<tr><td>The Netherlands National Trial Register</td><td>NTR</td><td>NTR[\s -]{0,1}[0-9]{2,4}</td><td>(?<![A-z])NTR</td></tr>
<tr><td>The Netherlands National Trial Register</td><td>NTR</td><td>NL[0-9]{3,4}</td><td>(?<![A-z])NTR</td></tr>
<tr><td>Pan African Clinical Trial Registry</td><td>PACTR</td><td>PACTR\s{0,1}[0-9]{10,}</td><td>PACTR</td></tr>
<tr><td>Peruvian Clinical Trial Registry</td><td>REPEC</td><td>PER(-|[0-9]){7}</td><td>PER</td></tr>
<tr><td>Sri Lanka Clinical Trials Registry</td><td>SLCTR</td><td>SLCTR/[0-9]{4}/[0-9]{3}</td><td>SLCTR</td></tr>
<tr><td>University hospital Medical Information Network</td><td>JPRN</td><td>UMIN[0-9]{9}</td><td>UMIN</td></tr>
<tr><td>Japanese Medical Association</td><td>JMA</td><td>JMA(-|[A-z]|[0-9]){8}</td><td>JMA</td></tr>
</table>

| | | | |
|---|---|---|---|
| Japan Pharmaceutical Information Center) | JapicCTI | JapicCTI-[0-9]{6} | Japic |
| Chinese Clinical Trial Registry | ChiCTR | ChiCTR.{0,1}[A-z]{0,3}.{0,1}[0-9]{7,8} | ChiCTR |
| Chinese Clinical Trial Registry | ChiCTR | CHICTR.{0,1}[A-z]{0,3}.{0,1}[0-9]{7,8} | ChiCTR |
| Figshare | figshare | [0-9]{2}.[0-9]{4}/.{0,3}figshare.[0-9]{7,} | figshare |
| Dryad | Dryad | [0-9]{2}.[0-9]{4}/dryad.[[0-9][A-z]]{5,} | Dryad |
| Figshare | figshare | [0-9]{2}.[0-9]{4}/.{0,3}FIGSHARE.[0-9]{7,} | FIGSHARE |
| Hong Kong Clinical Trial Registry | HKUCTR | HKU{0,1}CTR-[0-9]{4} | HKCTR |

### Supplementary Appendix 4. Data extraction

From PubMed-Medline xml files we extracted the article title, PubMed ID, journal, abstract, Trial Registration Number when available, author list and affiliation, date of submission, date of publication. The corresponding full text was retrieved from the database or through the library at the University of Geneva.

The following data was retrieved from the WHO-ICTRP database for study with a TRN, and from full text for the others: planned sample size, disease or condition of the participants, participating countries, funding, date of registration, date of first enrolment, contact details, funding, and type of intervention (pharmacological or non-pharmacological). The type of intervention was determined independently by two authors (SC, DBG) following a list of predetermined categories (see eAppendix5), and disagreements were resolved by a third author (DM).

A study was considered industry-funded if any private company was involved in funding, appeared as sponsor, or was named explicitly in the published article acknowledgments. Impact factors of each journal and year were recovered from the Scimago Journal and Country Rank website(1). The information about the journal publicly following ICMJE recommendations at the date of publication was obtained from the list of journals reported on the ICMJE website (2). If the journal did not appear in the list, we conducted an online search to identify the journal's statement about following the ICJME recommendations and if a date of adoption was reported.

Information about countries of enrollment was dichotomized in a two-level factor variable: non-international if recruitment was done only in one country or international if more than one country of recruitment was described.

#### References

1. Scimago Journal & Country Rank [Internet]. [cited 2024 Mar 13]. Available from: https://www.scimagojr.com/index.php

2. ICMJE | Home [Internet]. [cited 2024 Mar 13]. Available from: https://www.icmje.org/

**Supplementary Appendix 5. Definitions of sub-group of trials grouped under non-pharmacological interventions**

| Sub-Group | Definition |
|---|---|
| Behavioural | Interventions designed to affect the actions that individuals take regarding their health. It involves using behaviour analytic techniques to change, reduce or modify conducts in people. |
| Biological treatments | Interventions assessing the effect of blood components or cell therapies. |
| Delivery of health care services | Delivery of services within any a health system where patients receive the treatment and supplies. |
| Device | Interventions involving any instrument, apparatus, implement, machine, appliance, implant, reagent for in vitro use, software, material or other similar or related article, intended by the manufacturer to be used, alone or in combination for a medical purpose. |
| Education | Interventions that involved providing teaching, instruction or pedagogy activities to patients or other populations. |
| Exercise therapy | Interventions that assess the effect of sports or any physical training. |
| Food/plants/supplements | Any food, herbal or dietary supplements that are not classified as a drug. |
| Procedure | Interventions that involve any practice of a health practitioner that assesses a combination of special skills or abilities and may require drugs, devices, or both. |
| Surgical | Invasive interventions that require an incision, cutting into the skin, to access body tissue, organs, or other internal parts. |
| Wellness and spa | Interventions that assess the effectiveness of thermal baths, spas or massage therapies. |
| Other | Other interventions that do not fit into the former categories |

**Supplementary Appendix 6. Survey sent to corresponding authors and/or trial contact authors of unregistered trials**

**Trial registration practices**

| **Study author** | **Article title** |
|---|---|
| To the best our knowledge, the clinical trials reported in your article has not been registered in a WHO associated registry. | ○ The clinical trial has not been registered<br>○ The clinical trial has been registered in a WHO associated registry<br>○ The clinical trial has been registered in another registry<br>○ The clinical trial registry started before 2005<br>○ The article does not report the findings of a clinical trial |
| Please specify the registration number of your trial | _ _ _ _ _ _ _ _ _ _ _ _ _ |
| Why was your study not registered | ☐ We were not aware of the registration requirement.<br>☐ We did not think registration was important.<br>☐ We did not have enough time.<br>☐ We knew it was a requirement but forgot to do it.<br>☐ We thought the registration requirement only applied to industry funded trials.<br>☐ We thought the registration requirement only applied to drug-related trials.<br>☐ We did not realize that our study meets the definition of a clinical trial.<br>☐ Other reasons |
| Please specify why you're your study was not registered | _ _ _ _ _ _ _ _ _ _ _ _ _ |
| Please tick any factors that would have helped you to register your study | ☐ Registration number is required on the ethics form<br>☐ Having ethics submission automatically uploaded a clinical trial registry<br>☐ Having registration on a checklist provided by ethics committee or other regulatory body/other<br>☐ Having it included in my organization's procedures manual<br>☐ Registration required by journals<br>☐ Other |
| **Please specify a factor that would have helped you to register your study** | |

### Supplementary Table 3. Characteristics of RCTs stratified by registration status

| Variable | Category | Not Registered No. (%) | Retrospective No. (%) | Prospective No. (%) | P value | % missing |
|---|---|---|---|---|---|---|
| **Number of studies** | | 130 | 323 | 640 | | |
| **Continent** | Europe | 39 (31.2) | 142 (45.2) | 215 (34.5) | <0.001 | 2.7 |
| | Asia | 64 (51.2) | 85 (27.1) | 114 (18.3) | | |
| | North America | 10 (8.0) | 38 (12.1) | 95 (15.2) | | |
| | Transcontinental | 1 (0.8) | 11 (3.5) | 114 (18.3) | | |
| | Oceania | 3 (2.4) | 10 (3.2) | 68 (10.9) | | |
| | South America | 5 (4.0) | 22 (7.0) | 16 (2.6) | | |
| | Africa | 3 (2.4) | 6 (1.9) | 2 (0.3) | | |
| **International** | No | 119 (95.2) | 287 (91.4) | 466 (74.7) | <0.001 | 2.7 |
| | Yes | 6 (4.8) | 27 ( 8.6) | 158 (25.3) | | |
| **Funding** | Non-industry | 58 (63.7) | 231 (71.7) | 343 (53.6) | <0.001 | 3.7 |
| | Industry | 33 (36.3) | 91 (28.3) | 297 (46.4) | | |
| Number of authors | | 7.35 (4.83) | 8.54 (5.43) | 11.25 (6.94) | <0.001 | 0.0 |
| **Sample Size** | | 64.00 [45.00, 115.00] | 90.00 [59.50, 191.00] | 148.00 [76.00, 325.25] | <0.001 | 0.0 |
| **ICMJE[a]** | 0 | 97 (74.6) | 201 (62.2) | 233 (36.4) | <0.001 | 0.0 |
| | 1 | 33 (25.4) | 122 (37.8) | 407 (63.6) | | |
| **Impact Factor** | | 2.25 [1.71, 3.34] | 2.54 [2.20, 4.67] | 4.52 [2.52, 7.60] | <0.001 | 0.1 |
| **Delay (median month [IQR])** | between registration and recruitment start | | -9.23 [-26.87, -3.23] | 1.30 [0.08, 3.63] | <0.001 | 0 |
| **Type of Intervention** | Pharmacological | 36 (27.7) | 100 (31.0) | 330 (51.6) | <0.001 | 0.0 |
| | Exercise therapy | 22 (16.9) | 58 (18.0) | 72 (11.2) | | |
| | Procedure | 28 (21.5) | 50 (15.5) | 71 (11.1) | | |
| | Delivery of health care services | 13 (10.0) | 30 (9.3) | 52 (8.1) | | |
| | Device | 5 (3.8) | 19 (5.9) | 27 (4.2) | | |
| | Food/plants/supplements | 5 (3.8) | 15 (4.6) | 28 (4.4) | | |
| | Education | 5 (3.8) | 16 (5.0) | 12 (1.9) | | |
| | Surgical | 1 ( 0.8) | 13 ( 4.0) | 12 (1.9) | | |
| | Wellness | 7 (5.4) | 11 ( 3.4) | 5 (0.8) | | |
| | Behavioural | 3 (2.3) | 7 (2.2) | 11 (1.7) | | |
| | Biological treatment | 2 (1.5) | 4 (1.2) | 10 (1.6) | | |
| | Lifestyle | 3 (2.3) | 0 (0.0) | 10 (1.6) | | |
| **Condition** | Osteoarthritis | 20 (15.4) | 71 (22.0) | 136 (21.2) | 0.003 | 0.0 |
| | Rheumatoid arthritis | 36 (27.7) | 62 (19.2) | 124 (19.4) | | |
| | Spondylarthritis | 11 8.5) | 12 (3.7) | 58 (9.1) | | |
| | Connective tissue disease | 8 ( 6.2) | 24 ( 7.4) | 64 (10.0) | | |
| | Fibromyalgia | 12 (9.2) | 19 (5.9) | 24 (3.8) | | |
| | Arthroplasty | 1 (0.8) | 17 (5.3) | 29 (4.5) | | |
| | Low back pain | 6 (4.6) | 16 (5.0) | 24 (3.8) | | |
| | Gout | 0 (0.0) | 6 (1.9) | 18 (2.8) | | |
| | Fractures | 0 (0.0) | 9 (2.8) | 10 (1.6) | | |
| | Multiple diseases | 2 (1.5) | 4 (1.2) | 13 (2.0) | | |
| | Juvenile idiopathic arthritis | 4 (3.1) | 4 (1.2) | 10 (1.6) | | |
| | Knee pain | 1 (0.8) | 1 (0.3) | 12 (1.9) | | |
| | Anterior cruciate ligament | 2 (1.5) | 4 (1.2) | 6 (0.9) | | |
| | Neck pain | 3 (2.3) | 1 (0.3) | 8 (1.2) | | |
| | Subacromial impingement syndrome | 2 (1.5) | 5 (1.5) | 4 (0.6) | | |
| | Other | 22 (16.9) | 68 (21.1) | 100 (15.6) | | |
| **Journal** | BMC musculoskeletal | 7 (5.4) | 103 (31.9) | 103 (16.1) | <0.001 | 0.0 |

| Variable | Category | Not Registered No. (%) | Retrospective No. (%) | Prospective No. (%) | P value | % missing |
|---|---|---|---|---|---|---|
| | Disorders | | | | | |
| | Annals of the Rheumatic Diseases | 7 (5.4) | 37 (11.5) | 109 (17.0) | <0.001 | 0.0 |
| | Arthritis & Rheumatology | 1 (0.8) | 8 (2.5) | 58 (9.1) | | |
| | Osteoarthritis and Cartilage | 2 (1.5) | 17 (5.3) | 42 (6.6) | | |
| | Arthritis Care & Research | 6 (4.6) | 19 (5.9) | 33 (5.2) | | |
| | Clinical Rheumatology | 22 (16.9) | 20 (6.2) | 14 (2.2) | | |
| | The Lancet | 1 (0.8) | 6 (1.9) | 47 (7.3) | | |
| | Arthritis Research & Therapy | 1 (0.8) | 12 (3.7) | 36 (5.6) | | |
| | Rheumatology | 6 (4.6) | 13 (4.0) | 21 (3.3) | | |
| | Rheumatology International | 19 (14.6) | 11 (3.4) | 6 (0.9) | | |
| | JAMA | 0 (0.0) | 2 (0.6) | 28 (4.4) | | |
| | The Journal of Rheumatology | 8 (6.2) | 10 (3.1) | 12 (1.9) | | |
| | Annals of Internal Medicine | 0 (0.0) | 4 (1.2) | 20 (3.1) | | |
| | Arthritis and Rheumatism | 2 (1.5) | 9 (2.8) | 12 (1.9) | | |
| | International Journal of Rheumatic Diseases | 7 (5.4) | 5 (1.5) | 8 (1.2) | | |
| | Other | 41 (31.5) | 47 (14.6) | 91 14.2) | | |

**Abbreviations**

a. *ICMJE. International Committee of Medical Journals Editors*

**Supplementary Table 4. Proportion of RCTs published per year Prospective, Retrospective, or Unregistered, Stratified by ICMJE-compliant Journals**

| Year/type of registration | Overall No. (%) | Not ICMJE No. (%) | ICMJE No. (%) |
|---|---|---|---|
| 2009 - 2022 | 1093 | 531 | 562 |
| Not registered | 130 (11.9) | 97 (18.3) | 33 (5.9) |
| Retrospective | 323 (29.6) | 201 (37.9) | 122 (21.7) |
| Prospective | 640 (58.6) | 233 (43.9) | 407 (72.4) |
| 2009 | 15 | 9 | 6 |
| Not registered | 8 (53.3) | 6 (66.7) | 2 (33.3) |
| Retrospective | 5 (33.3) | 2 (22.2) | 3 (50.0) |
| Prospective | 2 (13.3) | 1 (11.1) | 1 (16.7) |
| 2010 | 32 | 17 | 15 |
| Not registered | 10 (31.2) | 7 (41.2) | 3 (20.0) |
| Retrospective | 11 (34.4) | 7 (41.2) | 4 (26.7) |
| Prospective | 11 (34.4) | 3 (17.6) | 8 (53.3) |
| 2011 | 45 | 24 | 21 |
| Not registered | 10 (22.2) | 9 (37.5) | 1 (4.8) |
| Retrospective | 19 (42.2) | 10 (41.7) | 9 (42.9) |
| Prospective | 16 (35.6) | 5 (20.8) | 11 (52.4) |
| 2012 | 49 | 30 | 19 |
| Not registered | 10 (20.4) | 9 (30.0) | 1 (5.3) |
| Retrospective | 17 (34.7) | 10 (33.3) | 7 (36.8) |
| Prospective | 22 (44.9) | 11 (36.7) | 11 (57.9) |
| 2013 | 57 | 25 | 32 |
| Not registered | 12 (21.1) | 10 (40.0) | 2 (6.2) |
| Retrospective | 15 (26.3) | 8 (32.0) | 7 (21.9) |
| Prospective | 30 (52.6) | 7 (28.0) | 23 (71.9) |
| 2014 | 66 | 23 | 43 |
| Not registered | 10 (15.2) | 7 (30.4) | 3 (7.0) |
| Retrospective | 25 (37.9) | 8 (34.8) | 17 (39.5) |
| Prospective | 31 (47.0) | 8 (34.8) | 23 (53.5) |
| 2015 | 67 | 33 | 34 |
| Not registered | 5 (7.5) | 4 (12.1) | 1 (2.9) |
| Retrospective | 18 (26.9) | 12 (36.4) | 6 (17.6) |
| Prospective | 44 (65.7) | 17 (51.5) | 27 (79.4) |

| Year/type of registration | Overall No. (%) | Not ICMJE No. (%) | ICMJE No. (%) |
|---|---|---|---|
| 2016 | 81 | 50 | 31 |
| Not registered | 8 (9.9) | 8 (16.0) | 0 (0.0) |
| Retrospective | 33 (40.7) | 20 (40.0) | 13 (41.9) |
| Prospective | 40 (49.4) | 22 (44.0) | 18 (58.1) |
| 2017 | 98 | 39 | 59 |
| Not registered | 10 (10.2) | 8 (20.5) | 2 (3.4) |
| Retrospective | 33 (33.7) | 16 (41.0) | 17 (28.8) |
| Prospective | 55 (56.1) | 15 (38.5) | 40 (67.8) |
| 2018 | 71 | 49 | 22 |
| Not registered | 12 (16.9) | 11 (22.4) | 1 (4.5) |
| Retrospective | 23 (32.4) | 20 (40.8) | 3 (13.6) |
| Prospective | 36 (50.7) | 18 (36.7) | 18 (81.8) |
| 2019 | 103 | 39 | 64 |
| Not registered | 8 (7.8) | 4 (10.3) | 4 (6.2) |
| Retrospective | 27 (26.2) | 16 (41.0) | 11 (17.2) |
| Prospective | 68 (66.0) | 19 (48.7) | 49 (76.6) |
| 2020 | 97 | 32 | 65 |
| Not registered | 6 (6.2) | 3 (9.4) | 3 (4.6) |
| Retrospective | 20 (20.6) | 10 (31.2) | 10 (15.4) |
| Prospective | 71 (73.2) | 19 (59.4) | 52 (80.0) |
| 2021 | 159 | 83 | 76 |
| Not registered | 9 (5.7) | 7 (8.4) | 2 (2.6) |
| Retrospective | 48 (30.2) | 37 (44.6) | 11 (14.5) |
| Prospective | 102 (64.2) | 39 (47.0) | 63 (82.9) |
| 2022 | 153 | 78 | 75 |
| Not registered | 12 (7.8) | 4 (5.1) | 8 (10.7) |
| Retrospective | 29 (19.0) | 25 (32.1) | 4 (5.3) |
| Prospective | 112 (73.2) | 49 (62.8) | 63 (84.0) |

**Supplementary Figure 1. Logit of the proportion of RCT with prospective registration as a function of impact factor (A) and of sample size (B). Errors bars correspond to the logit of the 95% confidence interval calculated with the Wilsons core interval**

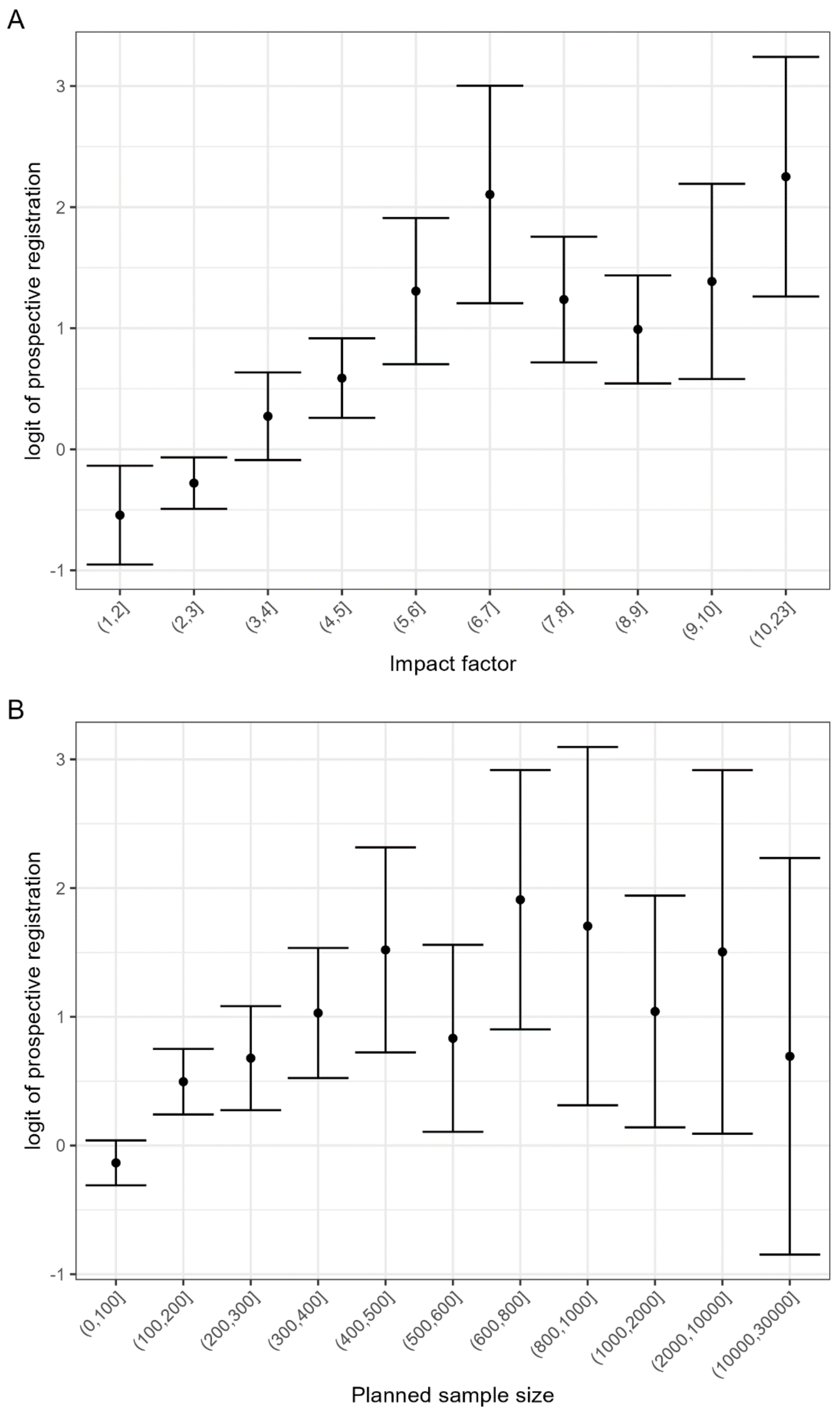

**Supplementary Table 5. Multivariable analysis considering the sub-group of interventions among non-pharmacological trials[a]**

| Variable | OR [95% CI] | P value |
|---|---|---|
| **International Trial** | | |
| No | Reference | |
| Yes | 1.94 [1.18, 3.18] | 0.009 |
| **Funding** | | |
| Non-Industry | Reference | |
| Industry | 1.01 [0.71, 1.44] | 0.96 |
| **Year** | | |
| | 1.16 [1.11, 1.20] | <0.001 |
| **Number of authors** | | |
| | 1.01 [0.98, 1.03] | 0.61 |
| **Journal endorsed ICMJE[b] recommendations** | | |
| No | Reference | |
| Yes | 1.22 [0.80, 1.85] | 0.36 |
| **Impact factor if < 5.5** | | |
| | 1.35 [1.15, 1.58] | <0.001 |
| **Impact factor if > 5.5** | | |
| | 1.04 [0.71, 1.51] | |
| **Sample size if < 500, per 100 participants** | | |
| | 1.24 [1.11, 1.38] | <0.001 |
| **Sample size if > 500, per 100 participants** | | |
| | 0.99 [0.79, 1.24] | |
| **Intervention** | | |
| Pharmacological | Reference | |
| Behavioural | 0.78 [0.29, 2.10] | 0.62 |
| Biological | 0.94 [0.30, 2.93] | 0.92 |
| Delivery of health care services | 0.57 [0.34, 0.97] | 0.04 |
| Device | 0.96 [0.50, 1.85] | 0.90 |
| Education | 0.34 [0.14, 0.79] | 0.01 |
| Exercise therapy | 0.75 [0.47, 1.19] | 0.22 |
| Food/plants/supplements | 1.10 [0.56, 2.16] | 0.78 |
| lifestyle | 1.88 [0.45, 7.84] | 0.38 |
| procedure | 0.73 [0.47, 1.15] | 0.18 |

| **Variable** | **OR [95% CI]** | **P value** |
|---|---|---|
| surgical | 0.51 [0.20, 1.27] | 0.15 |
| wellness | 0.24 [0.08, 0.70] | 0.00 |

a. Odds ratios (OR) and 95% confidence interval for each predictor of prospective registration obtained in the logistic regression analysis.
b. *ICMJE. International Committee of Medical Journals Editors*

**Supplementary Table 6.Multivariable analysis considering the conditions assessed in the included trials[a]**

| Variable | OR [95% CI] | P value |
|---|---|---|
| **International Trial** | | |
| No | Reference | |
| Yes | 1.94 [1.18, 3.18] | 0.009 |
| **Funding** | | |
| Non-Industry | Reference | |
| Industry | 1.01 [0.71, 1.44] | 0.96 |
| **Year** | | |
| | 1.16 [1.11, 1.20] | <0.001 |
| **Number of authors** | | |
| | 1.01 [0.98, 1.03] | 0.61 |
| **Journal endorsed ICMJE[b] recommendations** | | |
| No | Reference | |
| Yes | 1.22 [0.80, 1.85] | 0.36 |
| **Intervention** | | |
| Non-pharmacological | Reference | |
| Pharmacological | 1.67 [1.16, 2.39] | 0.06 |
| **Impact factor if < 5.5** | | |
| | 1.35 [1.15, 1.58] | <0.001 |
| **Impact factor if > 5.5** | | |
| | 1.04 [0.71, 1.51] | |
| **Sample size if < 500, per 100 participants** | | |
| | 1.24 [1.11, 1.38] | <0.001 |
| **Sample size if > 500, per 100 participants** | | |
| | 0.99 [0.79, 1.24] | |
| **Condition of interest/ population** | | |
| Rheumatoid Arthritis | Reference | |
| Anterior cruciate ligament | 2.60 [0.71, 9.48] | 0.15 |
| Arthroplasty | 3.11 [1.51, 6.39] | 0.002 |
| Connective tissue disease | 1.86 [1.03, 3.36] | 0.14 |
| Fibromyalgia | 1.63 [0.83, 3.21] | 0.15 |
| Fractures | 3.51 [1.27, 9.70] | 0.02 |
| Gout | 2.67 [0.94, 7.61] | 0.07 |

| Variable | OR [95% CI] | P value |
|---|---|---|
| Juvenile idiopathic arthritis | 1.03 [0.33, 3.24] | 0.95 |
| Knee pain | 14.14 [2.74, 73.06] | 0.002 |
| Low back pain | 3.56 [1.69, 7.50] | <0.001 |
| Multiple diseases | 1.72 [0.55, 5.34] | 0.35 |
| Neck pain | 8.01 [2.06, 31.05] | 0.003 |
| Osteoarthritis | 1.99 [1.26, 3.15] | 0.003 |
| Spondylarthritis | 1.31 [0.68, 2.54] | 0.42 |
| Subacromial impingement syndrome | 0.79 [0.18, 3.42] | 0.75 |
| Other | 1.82 [1.13, 2.93] | 0.01 |

a. Odds ratios (OR) and 95% confidence interval for each predictor of prospective registration obtained in the logistic regression analysis.
b. *ICMJE. International Committee of Medical Journals Editors*

**Supplementary Table 7. Characteristics of respondents and non-respondents that retrospectively registered RCTs and were invited to answer the study survey**

| Variable | Category | Overall No. (%) | Non-respondents No. (%) | Respondents No. (%) | p |
|---|---|---|---|---|---|
| | | 365 | 323 | 42 | |
| **Continent** | Europe | 162 (44.4) | 136 (42.1) | 26 (61.9) | 0.143 |
| | Asia | 104 (28.5) | 97 (30.0) | 7 (16.7) | |
| | South America | 30 (8.2) | 26 (8.0) | 4 (9.5) | |
| | North America | 28 (7.7) | 26 (8.0) | 2 (4.8) | |
| | Transcontinental | 22 (6.0) | 22 (6.8) | 0 (0.0) | |
| | Oceania | 11 (3.0) | 9 (2.8) | 2 (4.8) | |
| | Africa | 8 (2.2) | 7 (2.2) | 1 (2.4) | |
| **International** | No | 328 (89.9) | 286 (88.5) | 42 (100.0) | 0.041 |
| | Yes | 37 (10.1) | 37 (11.5) | 0 (0.0) | |
| **Sponsor** | Non-industry | 265 (72.6) | 230 (71.2) | 35 (83.3) | 0.141 |
| | Industry | 100 (27.4) | 93 (28.8) | 7 (16.7) | |
| **Number of authors mean (SD)** | | 8.85 (5.87) | 8.77 (5.50) | 9.48 (8.24) | 0.463 |
| **Target Sample Size median [IQR]** | | 90.00 [60.00, 190.00] | 90.00 [60.00, 190.00] | 101.50 [52.50, 196.75] | 0.875 |
| **ICMJE[a]** | No | 245 (67.1) | 223 (69.0) | 22 (52.4) | 0.047 |
| | Yes | 120 (32.9) | 100 (31.0) | 20 (47.6) | |
| **Impact Factor median [IQR]** | | 2.48 [2.18, 4.46] | 2.48 [2.17, 4.41] | 3.49 [2.23, 5.04] | 0.270 |
| **Type of Intervention** | Pharmacological | 105 (28.8) | 93 (28.8) | 12 (28.6) | 0.577 |
| | Procedure | 72 (19.7) | 67 (20.7) | 5 (11.9) | |
| | Exercise therapy | 62 (17.0) | 54 (16.7) | 8 (19.0) | |
| | Delivery of health care services | 41 (11.2) | 35 (10.8) | 6 (14.3) | |
| | Device | 23 (6.3) | 18 (5.6) | 5 (11.9) | |
| | Education | 19 (5.2) | 16 (5.0) | 3 (7.1) | |
| | Surgical | 16 (4.4) | 15 (4.6) | 1 (2.4) | |
| | Food/plants/supplements | 12 (3.3) | 12 (3.7) | 0 (0.0) | |
| | Biological treatment | 6 (1.6) | 6 (1.9) | 0 (0.0) | |
| | Behavioural | 5 (1.4) | 4 (1.2) | 1 (2.4) | |
| | Wellness | 4 (1.1) | 3 (0.9) | 1 (2.4) | |
| | Osteoarthritis | 79 (21.6) | 74 (22.9) | 5 (11.9) | 0.006 |
| **Condition** | Rheumatoid arthritis | 61 (16.7) | 53 (16.4) | 8 (19.0) | |
| | Arthroplasty | 24 (6.6) | 23 (7.1) | 1 (2.4) | |
| | Connective tissue disease | 32 (8.8) | 29 (9.0) | 3 (7.1) | |
| | Low back pain | 22 (6.0) | 21 (6.5) | 1 (2.4) | |
| | Fibromyalgia | 17 (4.7) | 16 (5.0) | 1 (2.4) | |
| | Fractures | 13 (3.6) | 13 (4.0) | 0 (0.0) | |
| | Spondylarthritis | 12 (3.3) | 9 (2.8) | 3 (7.1) | |
| | Multiple diseases | 6 (1.6) | 5 (1.5) | 1 (2.4) | |
| | Subacromial impingement syndrome | 6 (1.6) | 4 (1.2) | 2 (4.8) | |
| | Gout | 5 (1.4) | 5 (1.5) | 0 (0.0) | |
| | Anterior cruciate ligament | 4 (1.1) | 3 (0.9) | 1 (2.4) | |
| | Juvenile idiopathic arthritis | 3 (0.8) | 2 (0.6) | 1 (2.4) | |
| | Knee pain | 1 (0.3) | 0 (0.0) | 1 (2.4) | |
| | Neck pain | 1 (0.3) | 0 (0.0) | 1 (2.4) | |
| | Other | 79 (21.6) | 66 (20.4) | 13 (31.0) | |
| **Journal** | BMC musculoskeletal Disorders | 128 (35.1) | 116 (35.9) | 12 (28.6) | 0.341 |
| | Annals of the Rheumatic Diseases | 38 (10.4) | 33 (10.2) | 5 (11.9) | |

| Variable | Category | Overall<br>No. (%) | Non-respondents<br>No. (%) | Respondents<br>No. (%) | p |
|---|---|---|---|---|---|
| | Osteoarthritis and Cartilage | 21 (5.8) | 20 (6.2) | 1 (2.4) | |
| | Clinical Rheumatology | 19 (5.2) | 16 (5.0) | 3 (7.1) | |
| | Arthritis Research & Therapy | 15 (4.1) | 15 (4.6) | 0 (0.0) | |
| | Rheumatology | 14 (3.8) | 12 (3.7) | 2 (4.8) | |
| | Arthritis Care & Research | 12 (3.3) | 7 (2.2) | 5 (11.9) | |
| | Rheumatology International | 12 (3.3) | 11 (3.4) | 1 (2.4) | |
| | The Journal of Rheumatology | 12 (3.3) | 12 (3.7) | 0 (0.0) | |
| | The Lancet | 10 (2.7) | 8 (2.5) | 2 (4.8) | |
| | Advances in Rheumatology | 8 (2.2) | 7 (2.2) | 1 (2.4) | |
| | Arthritis & Rheumatology | 7 (1.9) | 7 (2.2) | 0 (0.0) | |
| | Clinical and Experimental Rheumatology | 7 (1.9) | 7 (2.2) | 0 (0.0) | |
| | Joint Bone Spine | 7 (1.9) | 6 (1.9) | 1 (2.4) | |
| | JAMA | 6 (1.6) | 4 (1.2) | 2 (4.8) | |
| | Other | 107 (29.3) | 95 (29.4) | 12 (28.6) | |

Abbreviations

*a- ICMJE. International Committee of Medical Journals Editors*

**Supplementary Table 8. Characteristics respondents and non-respondents that did not register RCTs and were invited to answer the study survey**

| Variable | Category | Overall No. (%) | Non-respondents No. (%) | Respondents No. (%) | p |
|---|---|---|---|---|---|
| | | 126 | 112 | 14 | |
| **Continent** | Asia | 64 (50.8) | 58 (51.8) | 6 (42.9) | 0.452 |
| | Europe | 41 (32.5) | 33 (29.5) | 8 (57.1) | |
| | North America | 9 (7.1) | 9 (8.0) | 0 (0.0) | |
| | South America | 5 (4.0) | 5 (4.5) | 0 (0.0) | |
| | Africa | 3 (2.4) | 3 (2.7) | 0 (0.0) | |
| | Oceania | 3 (2.4) | 3 (2.7) | 0 (0.0) | |
| | Transcontinental | 1 (0.8) | 1 (0.9) | 0 (0.0) | |
| **International** | No | 120 (95.2) | 107 (95.5) | 13 (92.9) | 1.000 |
| | Yes | 6 (4.8) | 5 (4.5) | 1 (7.1) | |
| **Funding** | Non-industry | 57 (64.0) | 50 (65.8) | 7 (53.8) | 0.605 |
| | Industry | 32 (36.0) | 26 (34.2) | 6 (46.2) | |
| **Number of authors mean (SD)** | | 7.32 (4.74) | 7.46 (4.92) | 6.21 (2.78) | 0.358 |
| **Target Sample Size median [IQR]** | | 64.50 [45.00, 111.50] | 64.00 [45.00, 110.50] | 76.50 [58.50, 140.00] | 0.330 |
| **ICMJE[a]** | No | 92 (73.0) | 85 (75.9) | 7 (50.0) | 0.082 |
| | Yes | 34 (27.0) | 27 (24.1) | 7 (50.0) | |
| **Impact Factor median [IQR]** | | 2.25 [1.77, 3.34] | 2.18 [1.77, 2.98] | 3.65 [1.55, 6.80] | 0.266 |
| **Type of Intervention** | Pharmacological | 36 (28.6) | 31 (27.7) | 5 (35.7) | 0.228 |
| | Procedure | 26 (20.6) | 24 (21.4) | 2 (14.3) | |
| | Exercise therapy | 20 (15.9) | 19 (17.0) | 1 (7.1) | |
| | Delivery of health care services | 13 (10.3) | 8 (7.1) | 5 (35.7) | |
| | Wellness | 8 (6.3) | 7 (6.2) | 1 (7.1) | |
| | Device | 5 (4.0) | 5 (4.5) | 0 (0.0) | |
| | Education | 5 (4.0) | 5 (4.5) | 0 (0.0) | |
| | Food/plants/supplements | 4 (3.2) | 4 (3.6) | 0 (0.0) | |
| | Behavioural | 3 (2.4) | 3 (2.7) | 0 (0.0) | |
| | Lifestyle | 3 (2.4) | 3 (2.7) | 0 (0.0) | |
| | Biological treatment | 2 (1.6) | 2 (1.8) | 0 (0.0) | |
| | Surgical | 1 (0.8) | 1 (0.9) | 0 (0.0) | |
| **Condition** | Rheumatoid arthritis | 33 (26.2) | 28 (25.0) | 5 (35.7) | 0.874 |
| | Osteoarthritis | 20 (15.9) | 19 (17.0) | 1 (7.1) | |
| | Spondylarthritis | 12 (9.5) | 11 (9.8) | 1 (7.1) | |
| | Fibromyalgia | 11 (8.7) | 10 (8.9) | 1 (7.1) | |
| | Low back pain | 6 (4.8) | 5 (4.5) | 1 (7.1) | |
| | Connective tissue disease | 8 (6.3) | 7 (6.2) | 1 (7.1) | |
| | Juvenile idiopathic arthritis | 4 (3.2) | 4 (3.6) | 0 (0.0) | |
| | Multiple diseases | 3 (2.4) | 2 (1.8) | 1 (7.1) | |
| | Anterior cruciate ligament | 2 (1.6) | 2 (1.8) | 0 (0.0) | |
| | Neck pain | 2 (1.6) | 2 (1.8) | 0 (0.0) | |
| | Subacromial impingement syndrome | 2 (1.6) | 1 (0.9) | 1 (7.1) | |
| | Arthroplasty | 1 (0.8) | 1 (0.9) | 0 (0.0) | |
| | Knee pain | 1 (0.8) | 1 (0.9) | 0 (0.0) | |
| | Other | 21 (16.7) | 19 (17.0) | 2 (14.3) | |
| **Journal** | Clinical Rheumatology | 22 (17.5) | 20 (17.9) | 2 (14.3) | |
| | Rheumatology | 19 (15.1) | 18 (16.1) | 1 (7.1) | 0.083 |

| Variable | Category | Overall No. (%) | Non-respondents No. (%) | Respondents No. (%) | p |
|---|---|---|---|---|---|
| | International | | | | |
| | Annals of the Rheumatic Diseases | 9 (7.1) | 5 (4.5) | 4 (28.6) | |
| | Archives of Rheumatology | 7 (5.6) | 6 (5.4) | 1 (7.1) | |
| | BMC Musculoskeletal Disorders | 7 (5.6) | 7 (6.2) | 0 (0.0) | |
| | International Journal of Rheumatic Diseases | 7 (5.6) | 6 (5.4) | 1 (7.1) | |
| | The Journal of Rheumatology | 7 (5.6) | 7 (6.2) | 0 (0.0) | |
| | Arthritis Care & Research | 6 (4.8) | 6 (5.4) | 0 (0.0) | |
| | Rheumatology | 5 (4.0) | 5 (4.5) | 0 (0.0) | |
| | Clinical and Experimental Rheumatology | 4 (3.2) | 4 (3.6) | 0 (0.0) | |
| | Musculoskeletal care | 4 (3.2) | 2 (1.8) | 2 (14.3) | |
| | Scandinavian Journal of Rheumatology | 4 (3.2) | 4 (3.6) | 0 (0.0) | |
| | Modern Rheumatology | 3 (2.4) | 3 (2.7) | 0 (0.0) | |
| | RMD Open | 3 (2.4) | 1 (0.9) | 2 (14.3) | |
| | International Journal of Rheumatology | 2 (1.6) | 2 (1.8) | 0 (0.0) | |
| | Other | 55 (43.7) | 49 (43.8) | 6 (42.9) | |

Abbreviation

a. ICMJE. International Committee of Medical Journals Editors
